# Plasma immune signatures associate with neurodegeneration in frontotemporal lobar degeneration

**DOI:** 10.64898/2026.09.28.26364155

**Authors:** Meira van Schaik, Kei Onn Lai, Noah L Shapiro, Alexander Jäck, Julia Goddard, Harry Crook, Roosmarijn Frohn, P Simon Jones, Sean YW Tan, Owen Swann, Rachel Mulvaney, Amanda Heslegrave, Henrik Zetterberg, Matthias Brendel, Kamen A Tsvetanov, James B Rowe, Maura Malpetti

## Abstract

Frontotemporal lobar degeneration (FTLD) encompasses clinically and biologically heterogeneous syndromes that are unlikely to be captured by a single blood biomarker. Although plasma neurofilament light (NfL) chain reflects neuroaxonal injury, multiplex profiling may identify complementary inflammatory, vascular and proteostatic processes relevant to clinical phenotype and progression. We characterised plasma CNS-related and inflammatory proteomic signatures across FTLD-related clinical syndromes and tested their associations with neuroaxonal injury, clinical severity, regional brain atrophy and survival. Plasma samples from 177 participants with behavioural variant frontotemporal dementia (bvFTD), primary progressive aphasia (PPA), progressive supranuclear palsy (PSP) or corticobasal syndrome (CBS) and 67 healthy controls were analysed using the high-sensitivity nucleic acid-linked immuno-sandwich assay (NULISA) platform. We quantified 130 CNS-related and 250 inflammation-related proteins. Differential expression and multivariate classification analyses were complemented by hierarchical clustering, penalised regression and survival analyses. Canonical correlation analysis tested multivariate associations between disease-associated proteins and regional MRI brain volumes. Sixty-one CNS-related and 61 inflammation-related proteins differed between participants with FTLD-related syndromes and controls after false-discovery-rate correction. NfL was the strongest individual discriminator between all patients and controls, with an area under the receiver operating characteristic curve of 0.91. In exploratory full-cohort models, the multivariate CNS signature achieved an area under the curve of 0.93, compared with 0.82 for the multivariate inflammation signature. However, inflammatory proteins provided stronger discrimination for selected syndrome comparisons, including PPA versus PSP. Among inflammation-related proteins, SPP1 and CD276 showed the strongest positive associations with NfL. Higher CD276 and SPP1 were associated with increased mortality, whereas higher amyloid-β42 was associated with lower mortality. A multivariate protein signature weighted towards tumour necrosis factor and immune signalling was associated with lower regional brain volumes across predominantly frontotemporal and limbic regions (canonical correlation 0.43; permutation p=0.018).

Plasma multiplex proteomics identified complementary dimensions of neuroaxonal injury and immune dysregulation across frontotemporal lobar degeneration-related syndromes. CNS-related proteins, particularly NfL, predominantly captured case-control neurodegeneration, whereas inflammatory signatures provided additional information about syndrome heterogeneity, structural brain changes and survival. SPP1, CD276 and broader immune-related signatures are candidates for independent biomarker validation and mechanistic investigation.

## Introduction

Frontotemporal lobar degeneration (FTLD) comprises a heterogeneous group of neurodegenerative disorders presenting with progressive changes in behaviour, language and movement (1). Its major clinical syndromes include behavioural variant frontotemporal dementia (bvFTD), primary progressive aphasia (PPA), progressive supranuclear palsy (PSP) and corticobasal syndrome (CBS), but clinical phenotype only imperfectly predicts the underlying molecular pathology (1). Importantly, frontotemporal dementia is one of the leading causes of early-onset dementia although it can also occur in later life, resulting in substantial clinical and societal burden (2). The clinicopathological heterogeneity of FTLD syndromes limits diagnostic confidence and complicates prognosis, patient stratification and recruitment to molecularly targeted trials. Although neuroimaging and cerebrospinal fluid biomarkers can improve diagnostic evaluation, their cost, availability or invasiveness constrain repeated and large-scale use. Blood biomarkers therefore offer an attractive route to scalable biological assessment.

Plasma neurofilament light chain (NfL) is a sensitive marker of neuroaxonal injury in FTLD: it can help distinguish FTLD syndromes from primary psychiatric disorders and predicts phenoconversion and progression in familial disease (3,4). However, NfL is not specific to FTLD and primarily reflects the intensity of neuronal injury rather than the molecular processes producing it. FTLD pathogenesis involves interacting abnormalities in protein homeostasis, synaptic function, glial responses, vascular integrity and immunity, which are unlikely to be represented adequately by a single analyte. Multiplex proteomic profiling can measure these processes simultaneously, test whether combinations of proteins provide complementary diagnostic or prognostic information, and identify co-expression signatures that may reveal shared or syndrome-selective biology.

Inflammation is particularly relevant to this approach. Neuropathological, genetic and in vivo imaging studies implicate microglial activation, astrocytic responses and dysregulated immune signalling across FTLD-related disorders (5–8). In parallel, peripheral inflammatory profiles have been related to central microglial activation and survival across FTLD syndromes (9), suggesting that circulating proteins may provide clinically relevant information about neuroimmune activity. However, peripheral inflammatory proteins can reflect systemic responses, neurovascular signalling, consequences of tissue injury or compensatory regulation. Establishing their relevance requires convergence with independent measures of neurodegeneration, brain structure and clinical outcome. In this context, whether specific inflammatory pathways differ across syndromes and how they relate to neurodegeneration and progression remain poorly defined.

The nucleic acid-linked immuno-sandwich assay (NULISA) provides high-sensitivity, multiplex measurement of low-abundance plasma proteins, including established markers of neurodegeneration alongside proteins involved in inflammation, vascular biology, synaptic function and proteostasis (10). Prior studies have demonstrated the feasibility of NULISA profiling across neurodegenerative diseases (11–13) and in small cohorts of genetic frontotemporal dementia (14), but the relationship between broad CNS and inflammatory signatures, clinical heterogeneity, neuroaxonal injury, regional atrophy and prognosis across major FTLD-related syndromes remains incompletely defined.

In this study, we profiled plasma CNS-related and inflammatory proteins in participants with bvFTD, PPA, PSP and CBS, and in healthy controls. We first tested the ability of individual proteins and multivariate signatures to differentiate between groups. We then examined whether disease-associated proteins formed biologically coherent modules (“profiles”) and whether these proteins or profiles were associated with plasma NfL, clinical severity, survival and regional brain volumes. We hypothesised that CNS-related proteins would predominantly capture case-control neurodegeneration, whereas inflammatory signatures would provide complementary information about syndrome heterogeneity and disease progression and would covary with neuroaxonal injury and atrophy in FTLD-vulnerable networks. By investigating disease-relevant blood protein signatures, we aim to clarify disease mechanisms underlying FTLD and identify clinically relevant related biomarkers, which can support better prognosis and the identification of therapeutic targets.

## Material and methods

### Participants

Patient participants were recruited from specialised clinical centres for cognitive and movement disorders. Patient participants were included with a clinical diagnosis of FTLD-related syndromes according to the clinical consensus diagnostic criteria of bvFTD (n = 54) (15), PPA (n = 22; 13 nfvPPA, 7 svPPA, 2 mixed PPA) (16), PSP (n = 80) (17) and CBS (n = 21) (18). We also recruited healthy controls (n = 67). Exclusion criteria for recruitment of both patient participants and controls included concurrent major psychiatric illness or other severe physical illness, or a history of other significant neurological illness.

Participants underwent cognitive assessment using the Addenbrooke’s Cognitive Examination Revised (ACER, 0-100 scale), with lower scores indicating worse cognitive performance (19) and the Cambridge Behavioural Inventory, revised (CBIR, 0-180 scale), where higher scores indicate worse behavioural and functional symptoms (20). Participants with PSP and CBS also underwent clinical assessment using the Progressive Supranuclear Palsy Rating Scale (PSPRS 0-100 scale) (21). Higher PSPRS scores are indicative of worse disease severity. Participants with mental capacity provided written informed consent to take part in the study, in line with the Declaration of Helsinki. For participants who were incapacitated, consultee procedures were followed in accordance with UK law. The research protocols were approved by the National Research Ethics Service’s East of England Cambridge Central Committee (REC references 07/Q0102/3, 15/EE/0270, 24/EE/0214, and 07/H0307/64).

### Sample collection and processing

Blood samples were obtained by venipuncture and collected in ethylenediaminetetraacetic acid (EDTA) tubes. EDTA tubes were centrifuged to isolate plasma and stored at −70 °C until further analysis. Plasma samples from all participants were subsequently analysed using the NULISAseq assay. The NULISA CNS panel consists of 130 CNS-disease related fluid biomarkers, including markers of neurodegeneration (e.g. NfL), neuropathology (e.g. phosphorylated tau isoforms), inflammation and vascular health. The NULISA inflammation panel consists of 250 markers of inflammation and immune-mediated responses, including interleukins, chemokines and complement proteins (10).

Sample and data analysis was performed according to kit manufacturer protocols, which includes log2 transformation of the data and NULISA Protein Quantification (NPQ) on the logarithmic scale (Alamar Biosciences), as described previously (12).

### Statistical analysis

All analyses were performed using R (version 4.5.1).

Descriptive demographic and clinical variables were compared between groups using chi-square tests (sex), analysis of variance tests (age, ACER, CBIR), Welch’s t-tests (PSPRS) and Fisher’s exact test (survival rates). Post-hoc testing (Tukey’s HSD and pairwise Fisher’s exact tests with multiplicity adjustment) was applied to variables that showed significant differences across groups.

Across all analyses p values <0.05 were considered significant. P values were adjusted for multiple comparisons, using False Discovery Rate (FDR) correction.

### Differential expression analysis

Empirical Bayes–moderated linear models (limma R package) were applied to assess differential expression between FTLD and control and pairwise comparisons of groups. All limma analyses included age and sex as covariates. The CNS and inflammation panels were assessed separately. Results were visualised with volcano plots, and the top 10 most significant markers were labelled.

Next, receiver operating characteristic (ROC) analysis was performed to assess how well biomarkers selected via differential expression analysis could discriminate between groups. For each group comparison, the top 10 most significant biomarkers were selected for ROC analysis. Binary classification signatures were constructed for each of the comparisons (i.e. FTLD vs control and pairwise comparisons for all groups). Sensitivity and specificity values across classification thresholds were used to generate ROC curves, and area under the curve (AUC) values were calculated to quantify performance. Diagnostic performance was assessed independently for each of the top 10 markers, and markers were also combined by creating multivariate logistic regression models. The analysis was done separately for the CNS and inflammation panel. The initial ROC analysis was performed on the full dataset without resampling or cross-validation to explore performance. To assess how robust biomarker performance is and to reduce overfitting, a ROC analysis was performed using a bootstrap resampling approach. For each group comparison, 200 bootstrap samples were generated by randomly resampling participants from the original dataset, allowing participants to be selected multiple times (22). Within each bootstrap training sample, differential expression analysis was performed (using limma), and the top 10 most significant markers were selected. These markers were then used to create an elastic net regularised logistic regression model with 5-fold internal cross-validation. The performance of the model was assessed by testing on the out-of-bag participants that were not included in the bootstrap training samples. The predicted probabilities from these observations were combined across the bootstrap iterations to compute pooled ROC curves and AUC values. To assess biomarker stability, the frequency with which biomarkers were selected across the 200 bootstrap iterations was reported.

Differential expression analysis was also performed on the combined CNS and inflammation panels, which included 328 unique biomarkers after removing overlapping markers from the inflammation panel. The resulting biomarkers (significantly different between groups) were included in the next analyses with clinical severity scores, survival rates, neurodegeneration markers, and hierarchical clustering.

### Biomarker clustering and PCA

The CNS and inflammation biomarkers that were differentially expressed between FTLD and controls (FDR < 0.05) were selected for clustering analysis. Control participants were excluded from this analysis. To identify groups of co-expressed biomarkers, pairwise Pearson correlation coefficients were calculated across biomarkers, keeping only positive correlations. A biomarker dissimilarity matrix was generated, and hierarchical clustering was performed using Ward’s minimum variance method (Ward.D2). The optimal number of biomarker clusters was chosen using an elbow plot of within-cluster dissimilarity. Principal component analysis (PCA) was performed independently on each cluster of biomarkers. The first principal component (PC1) was selected as a score to summarise the cluster module’s expression for each participant. The sign of PC1 was inverted to orient the loadings predominantly in the positive direction to improve interpretability. The differences in PC1 scores between clinical syndromes were assessed per cluster module, including age and sex as covariates. The PC1 scores of the biomarker clusters were linked to survival via Cox regression analysis, including age, sex, and group as covariates. In addition, Spearman correlations between the PC1 cluster scores and NfL expression were computed to assess the link of cluster expression with neuroaxonal injury.

### Correlations with plasma NfL, clinical severity and mortality risk

First, Spearman correlations were computed between all biomarkers on the inflammation panel differentially expressed in FTLD and NfL. NfL expression functioned as a proxy measure of neuroaxonal injury.

Second, CNS and inflammation biomarkers that differed between patients and controls were related to three different clinical scores. The relationship between biomarkers and total ACER score (0-100), and total CBIR score (0–180) was assessed across all clinical syndromes. The associations between biomarkers and PSPRS (0-100) were only investigated in PSP and CBS groups. Elastic net regression (α=0.5) was used to preselect the best predictors, which were then used to create linear regression models with age, sex and group as covariates. The standardised β regression coefficients were reported for ease of interpretation.

Finally, CNS and inflammation biomarkers differentially expressed in patients versus controls were linked to mortality. The survival analysis was performed across all clinical syndromes. Penalised Cox regression with elastic net regularised regression was used to select biomarkers associated with mortality risk. The selected biomarkers were included in the final Cox proportional hazard regression model, including sex, age and group as covariates. Time to event was defined as time of blood sample collection to time of death or to the census date for participants alive at the time of the analysis (3 March 2026).

### Canonical correlation analysis

Canonical correlation analysis (CCA) was performed to assess multivariate associations between regional MRI brain volumes and expression of inflammation and CNS proteins that were differentially expressed in FTLD. Regional brain volumes were divided by total intracranial volume prior to CCA. Brain regions were defined according to the Hammersmith atlas. CCA was performed in MATLAB using the CSA toolbox. Prior to the CCA, protein and MRI values were z standardised. Regularisation parameters were set automatically by the algorithm. 10-fold cross-validation with 1000 repetitions was applied to improve model stability. Statistical significance was tested by running 1000 permutations per cross-validation partition, creating empirical null distributions against which the observed canonical correlations were compared. Next, CCA results were imported into R Studio. The direction of both the MRI and protein CCA loadings and subject scores was inverted to facilitate interpretation. Linear modelling was performed between MRI and protein subject-level scores with group as a covariate, including a variation with a protein score x group interaction term (23,24). Cox proportional hazard regression was performed in patient participants to assess associations between protein and MRI canonical scores and survival, including age and sex as covariates.

## Results

### Cohort characteristics

Demographics and clinical characteristics of the control and patient participants are summarised in Table 1. There were no significant differences in sex ratio across groups. Controls and participants with bvFTD were significantly younger than participants with PPA and PSP. Group-level testing showed a significant difference in ACER scores, although post-hoc testing was non-significant. There were no significant differences in PSPRS scores. Participants with bvFTD had significantly higher CBIR scores in comparison to participants with PPA, and participants with PPA showed a lower proportion of deaths at the census date compared to participants with PSP.

**Table 1:** Demographic and clinical information of all participants in NULISA plasma proteomic analysis.

| Group | N | Sex (F/M) | Age (mean ± sd) | ACER (mean ± sd) | PSPRS (mean ± sd) | CBIR (mean ± sd) | Proportion dead |
| --- | --- | --- | --- | --- | --- | --- | --- |
| Control | 67 | 28/39 | 68.1 ± 6 <sup>a</sup> | - | - | - | - |
| bvFTD | 54 | 19/35 | 65.6 ± 8.5 <sup>a</sup> | 70.6 ± 21.1 | - | 73.6 ± 27.6 <sup>b</sup> | 12/54 |
| PPA | 22 | 11/11 | 73.3 ± 7.7 | 68.6 ± 15.4 | - | 19.8 ± 34 | 0/22 <sup>c</sup> |
| PSP | 80 | 33/47 | 72.5 ± 7.8 | 79.5 ± 13.5 | 36.4 ± 12.7 | 46.1 ± 39.2 | 25/80 |
| CBS | 21 | 14/7 | 69.1 ± 7.7 | 85.2 ± 13.4 | 30.9 ± 22.2 | 64.4 ± 38.9 | 3/21 |
| Group comparison | | $\chi^2(4) = 6.72, p = 0.151$ | $F(4) = 8.95, p < 0.001$ | $F(3) = 3.40, p = 0.022$ | $t(10.8) = -0.74, p = 0.472$ | $F(3) = 3.49, p = 0.025$ | Fisher's exact test, $p = 0.006$ |

<sup>a</sup> $p < 0.05$ in comparison to PPA and PSP in post-hoc testing
<sup>b</sup> $p < 0.05$ in comparison to PPA in post-hoc testing
<sup>c</sup> $p < 0.05$ in comparison to PSP in post-hoc testing

### CNS and inflammatory plasma signatures distinguish FTLD-related syndromes from controls

Differential expression analysis revealed 61 CNS biomarkers with significant differences between the FTLD and control groups (FDR < 0.05), of which 37 were upregulated and 24 were downregulated in FTLD (Figure 1A). Among the 10 most significant biomarkers, upregulated biomarkers were associated with neuroaxonal injury (NfL), inflammation (SAA1, IL6, and IL2), tissue remodelling and proteostasis responses (POSTN and CST3), neuronal/neuroimmune signalling (TAFA5), and neuronal activity regulation (CALB2). The downregulated markers were related to synaptic plasticity and memory function (NRGN) and tissue repair and growth (FGF2).

**Figure 1.**
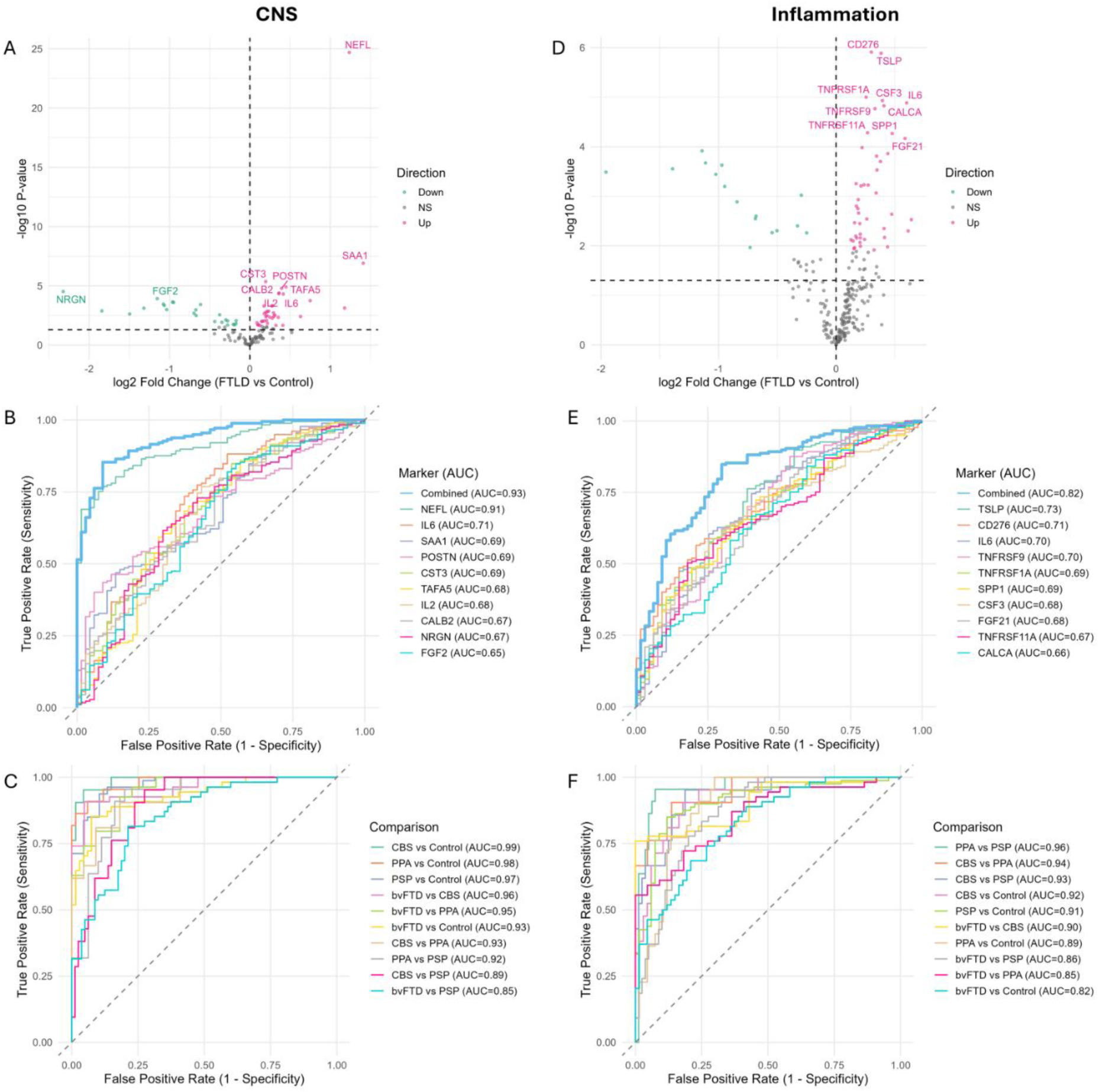
CNS-related and inflammatory plasma proteins distinguish FTLD-related clinical syndromes from controls. Volcano plots show differential expression of CNS-related (A) and inflammation-related proteins (D) in FTLD-related clinical syndromes versus controls, adjusted for age and sex. Labelled proteins are the 10 most significant after false-discovery-rate correction. Receiver-operating-characteristic curves show individual and combined top-10 protein models for CNS-related (B) and inflammation-related proteins (E), and combined models for pairwise group comparisons (C, F). Area under the curve values are shown. Internal validation is reported in Supplementary Figs. 1-2. Abbreviations: FTLD = frontotemporal lobar degeneration; CNS = central nervous system.

The diagnostic performance of the top 10 most significant differentially expressed markers was evaluated via ROC analysis (Figure 1B). Out of the individual markers, NfL showed the best performance in discriminating between FTLD and control with an AUC of 0.91, followed by IL6 (AUC = 0.71), and SAA1 (AUC = 0.69). The multivariable logistic regression model combining all 10 markers showed an AUC of 0.93, achieving slightly better discrimination than NfL alone.

Differential expression analysis of CNS biomarkers between groups showed significant differences between all pairwise group comparisons except for CBS versus PPA (Supplementary figure 3). The diagnostic performance of the multivariate model of the top 10 most significant CNS biomarkers for each group comparison is shown in Figure 1C. The best discrimination was achieved between CBS and the control group (AUC = 0.99), although all models showed strong discriminatory performance between groups, with a lowest AUC of 0.85 for discrimination between bvFTD and PSP.

The inflammation panel revealed 61 biomarkers with differential expression in FTLD compared to controls, with some overlap with the inflammatory markers included in the CNS panel. Out of the differentially expressed biomarkers, 45 were upregulated and 16 were downregulated in FTLD (Figure 1D). The top 10 most significant biomarkers were all upregulated. These biomarkers were related to immune regulation (CD276), pro-inflammatory immune activation (TSLP, IL6, CSF3, TNFRSF1A, TNFRSF9, TNFRSF11A), tissue remodelling and stress responses (SPP1, FGF21, CALCA).

TSLP showed the best discriminatory performance between FTLD and control among the top 10 most significant markers with an AUC = 0.73 (Figure 1E), followed by CD276 (AUC = 0.71), and IL6 (AUC = 0.70). The combined model performed better than the individual inflammation markers with an AUC of 0.82, although performance was still not as high as that of the combined CNS biomarker model.

The inflammation biomarkers showed significant differential expression in almost all pairwise group comparisons, apart from bvFTD versus PSP and CBS versus PPA (Supplementary figure 4). The multivariate inflammation biomarker models performed generally well in discriminating between groups (Figure 1F). The strongest discriminative performance was between PPA and PSP, with an AUC of 0.96. While the CNS panel was overall better at discriminating patient groups from the control group, the inflammation panel showed stronger performance for some of the syndrome vs syndrome comparisons.

### Co-expression modules across FTLD-syndromes capture immune, endothelial and proteostasis-related signatures

Differential expression analysis combining the inflammation and CNS panels identified 104 significant biomarkers, with 71 upregulated and 33 downregulated in FTLD (Supplementary figure 5A). Hierarchical clustering grouped these biomarkers of interest into biological modules by their expression patterns (Supplementary figure 5C). The number of clusters (k = 4) was selected by plotting within cluster dissimilarity against k number of clusters in an elbow plot (Supplementary figure 5B).

PCA was used to summarise the overall expression trends for each of the biomarker clusters. Cluster 1 comprised 26 markers, with the first principal component (PC1) explaining 22% of the variance. Cluster 2 was the smallest cluster, containing 13 markers, with PC1 explaining 30% of the variance. Cluster 3 contained 37 markers with PC1 accounting for 45% of the variance. Finally, cluster 4 consisted of 28 markers, with PC1 explaining 73% of the variance. Clusters were named based on the biological function of the proteins, prioritising those with the top 10 highest PC1 loadings (Figure 2A). Cluster 1 was a mix of biomarkers related to immune signalling, (CD276, CX3CL1, IL13RA2 and CCL11), endothelial activation (VCAM1, SLIT2), remodelling (POSTN and SLIT2) and tau pathology (pTau-217, BD-pTau-181, BD-pTau-217). Cluster 2 included biomarkers involved in systemic inflammation and acute immune activation (SAA1, IL6, CRP and IL17C), immune recruitment and regulation (IL1RN, CCL7, CCL13 and CSF3) and tissue regeneration (HGF and SMOC1). Cluster 3 was composed of biomarkers involved in TNF signalling (TNFRSF1B, TNFRSF4, TNFRSF11A, TNF, TNFRSF1A), T cell activation (CD83, IL15RA, LGALS9 and CTLA4), and proteostasis (CST3). Cluster 4 included markers related to cellular stress (MIF, IRAK4, CTF1), metabolism and homeostasis (PGK1 and ANXA5), and neuropathology (TARDBP, HTT, pSNCA.129, NRGN, Oligo.SNCA). The full lists of proteins in each cluster and their loadings can be found in Supplementary table 7.

**Figure 2.**
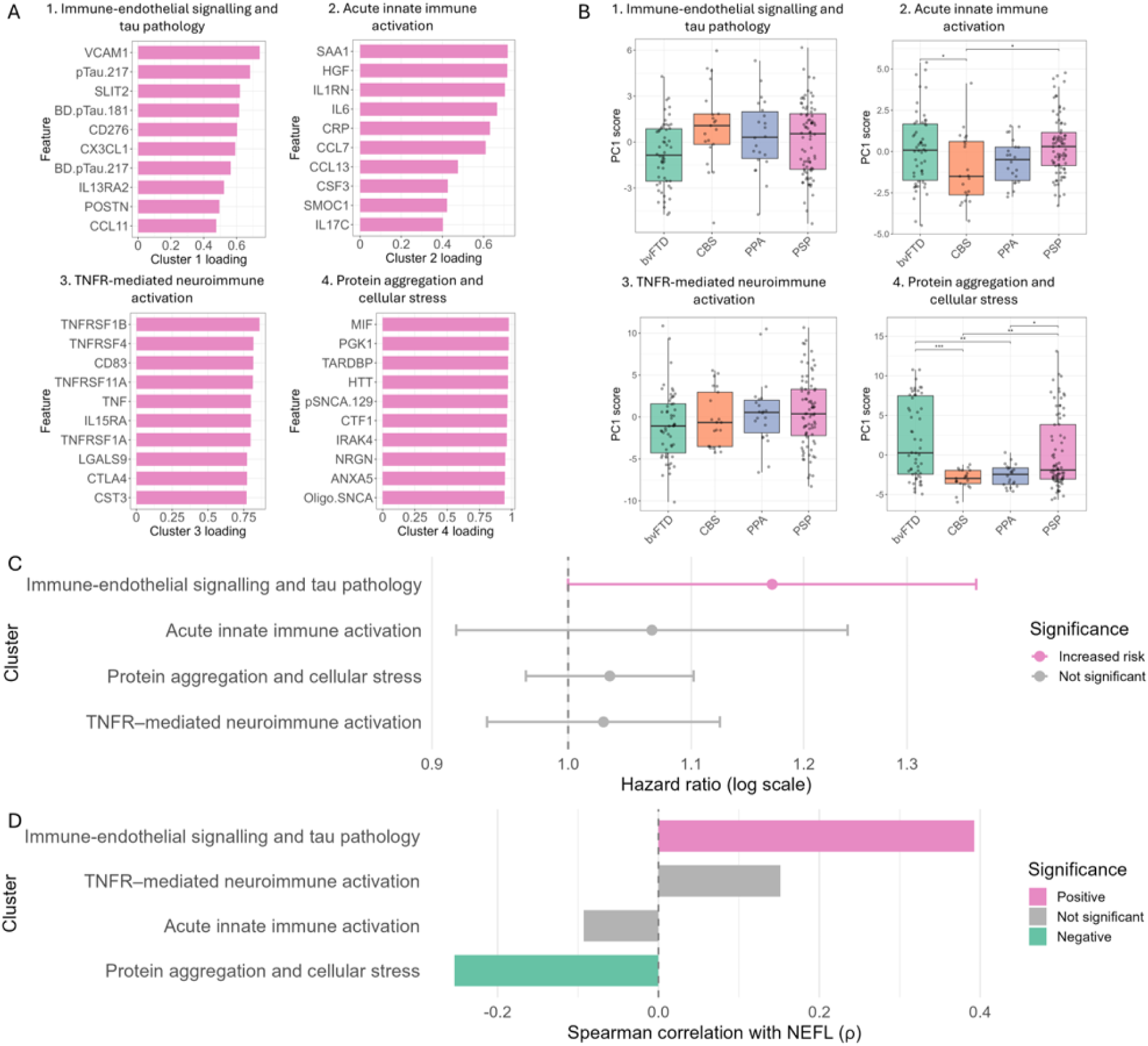
Co-expression modules of disease-associated plasma proteins across FTLD-related clinical syndromes. Hierarchical clustering of proteins differentially expressed in FTLD-related clinical syndromes versus controls identified four co-expression modules. The 10 proteins with the highest PC1 loadings are shown for each module (A). PC1 scores are compared across groups (B), related to survival in Cox regression models adjusted for age, sex and group (C), and correlated with plasma NfL (D). The cluster 1 association with survival did not meet the prespecified significance threshold (P=0.05). Abbreviations: PC1 = first principal component; NfL = neurofilament light chain.

PC1 scores of each cluster are shown per patient group (Figure 2B). Cluster 1 and 3 showed no significant differences across patient groups, while cluster 2 (acute innate immune activation) showed higher expression in bvFTD and PSP in comparison to CBS, and cluster 4 (protein aggregation and cellular stress) was expressed higher in bvFTD and PSP compared to both CBS and PPA.

Cox regression analysis revealed that cluster 1 (immune-endothelial signalling and tau pathology) was related to increased mortality risk (Figure 2C, p=0.05). Interestingly, cluster 1 was also positively correlated to NfL (Figure 2D), linking it with neuroaxonal injury. In contrast, cluster 4 was negatively correlated with NfL.

### Inflammatory proteins associate with neuroaxonal injury, mortality risk and clinical scores

Out of the 61 biomarkers from the inflammation panel that were differentially expressed in FTLD compared to control, 51 were significantly correlated with NfL (FDR < 0.05), a marker of neuroaxonal injury (Figure 3A). Of these 51 inflammation markers, 36 were positively correlated with NfL and 15 were negatively correlated. The top 10 most significant positive correlations with NfL included several markers that were also in the top 10 of the most significantly upregulated markers in FTLD (SPP1, CD276, TNFRSF9, TSLP, TNFRSF11A). CX3CL1 or fractalkine, a chemokine involved in neuroimmune interactions, was also among the strongest positive correlations with NfL. Among the strongest negative correlations were the growth factors EGF and FGF2, which engage in cell survival and tissue repair, and immune regulatory proteins CD40LG and MIF, which are related to adaptive immune signalling and inflammatory stress responses, respectively.

**Figure 3.**
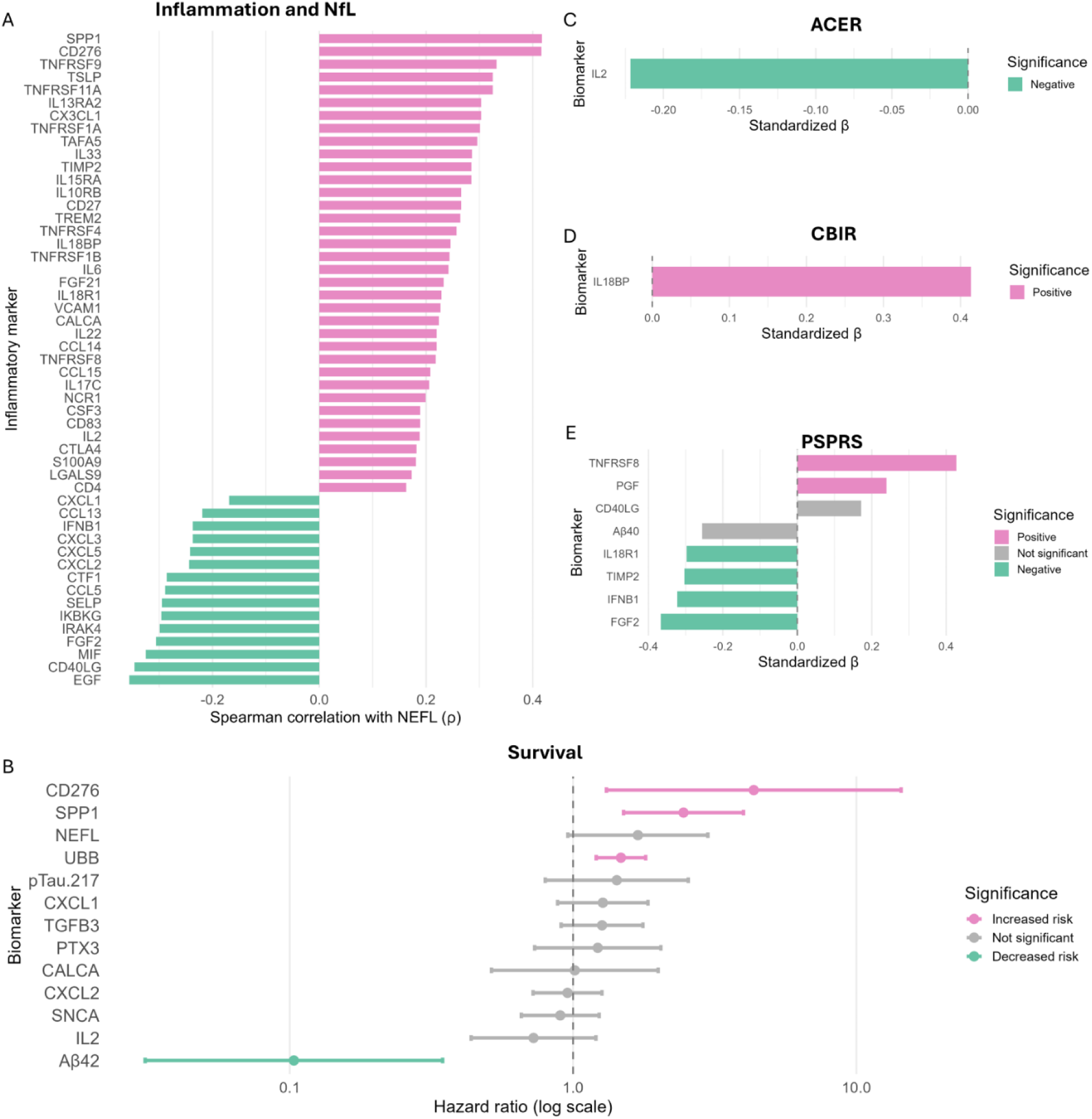
Plasma proteins associated with neuroaxonal injury, survival and clinical severity. Spearman correlations between NfL and inflammation-related proteins differentially expressed in FTLD-related clinical syndromes are shown in A. Proteins associated with survival after elastic-net selection and adjusted Cox regression are shown in B. Elastic-net-selected proteins associated with ACE-R (C), CBI-R (D) and PSPRS scores (E) are shown as standardised regression coefficients from models adjusted for age, sex and clinical group. Abbreviations: NfL = neurofilament light chain; ACE-R = Addenbrooke’s Cognitive Examination– Revised; CBI-R = Cambridge Behavioural Inventory–Revised; PSPRS = Progressive Supranuclear Palsy Rating Scale.

Among the differentially expressed inflammation and CNS biomarkers (Supplementary figure 5A), elastic net regression selected 13 biomarkers as the best predictors for survival, of which 4 showed significant associations with mortality risk (Figure 3B). CD276, SPP1, and UBB were linked to higher mortality risk. UBB encodes ubiquitin B, involved in protein degradation. Aβ42 was the only biomarker linked with lower mortality risk.

Biomarkers were linked to total clinical scores via elastic net regression selection. There were 86 participants with FTLD syndromes with ACER test scores available within 6 months of blood collection. Elastic net regression selected IL2 as the only biomarker associated with ACER. Linear modelling showed a negative association between IL2 and total ACER scores. (Figure 3C). Since high ACER scores are indicative of better cognitive abilities, this implies that high IL2 expression was related to worse cognitive outcomes. There were 43 patient participants with available CBIR scores within 6 months of blood collection. The elastic regression model selected IL18BP as being positively related to CBIR scores (Figure 3D). High CBIR scores indicate worse disease severity, meaning that high expression of IL18BP was related to more severe clinical symptoms. PSPRS scores were only assessed for participants with PSP and CBS. From these disease groups, 44 participants had PSPRS scores within 6 months of blood collection. The PSPRS model selected several different biomarkers that were significantly related to PSPRS scores (Figure 3E). TNFRSF8, a regulator of cell growth, and PGF, an endothelial marker, showed positive relationships with overall PSPRS scores, and were thus related to worse clinical performance. FGF2 (growth factor), IFNB (cytokine), TIMP2 (matrix metalloprotease inhibitor), and IL18R1 (IL18 receptor) were negatively related to PSPRS scores and were therefore linked with better clinical performance.

### A multivariate immune-related protein signature is associated with regional brain atrophy

Canonical correlation analysis (CCA) was performed on protein biomarkers differentially expressed in FTLD and regional brain volumes. The first canonical mode identified a significant association between protein biomarker expression and MRI measures of brain volumes (canonical correlation ρ = 0.43, permutation p = 0.018). Since the sign of the canonical variates is arbitrary, the sign of both the MRI and protein biomarker loadings and subject scores was inverted after CCA for improved interpretability. The significant positive correlation indicates that high expression of protein biomarkers with highly positive loadings is associated with smaller brain volumes in brain regions with highly negative loadings. Out of the 104 biomarkers in the CCA, 77 had positive canonical loadings and 27 had negative loadings (Figure 4A). Among the 10 protein biomarkers with the highest canonical loadings were markers associated with TNF signalling (TNFRSF1B, TNFRSF1A, TNFRSF11A and TNF), immune activation (TSLP, TREM1, CD83 and CD27), immune regulation (IL18BP) and tissue homeostasis (CST3). The negatively loaded proteins included several proteins that were also in cluster 4, which was negatively associated with NfL (Figure 2D)

**Figure 4.**
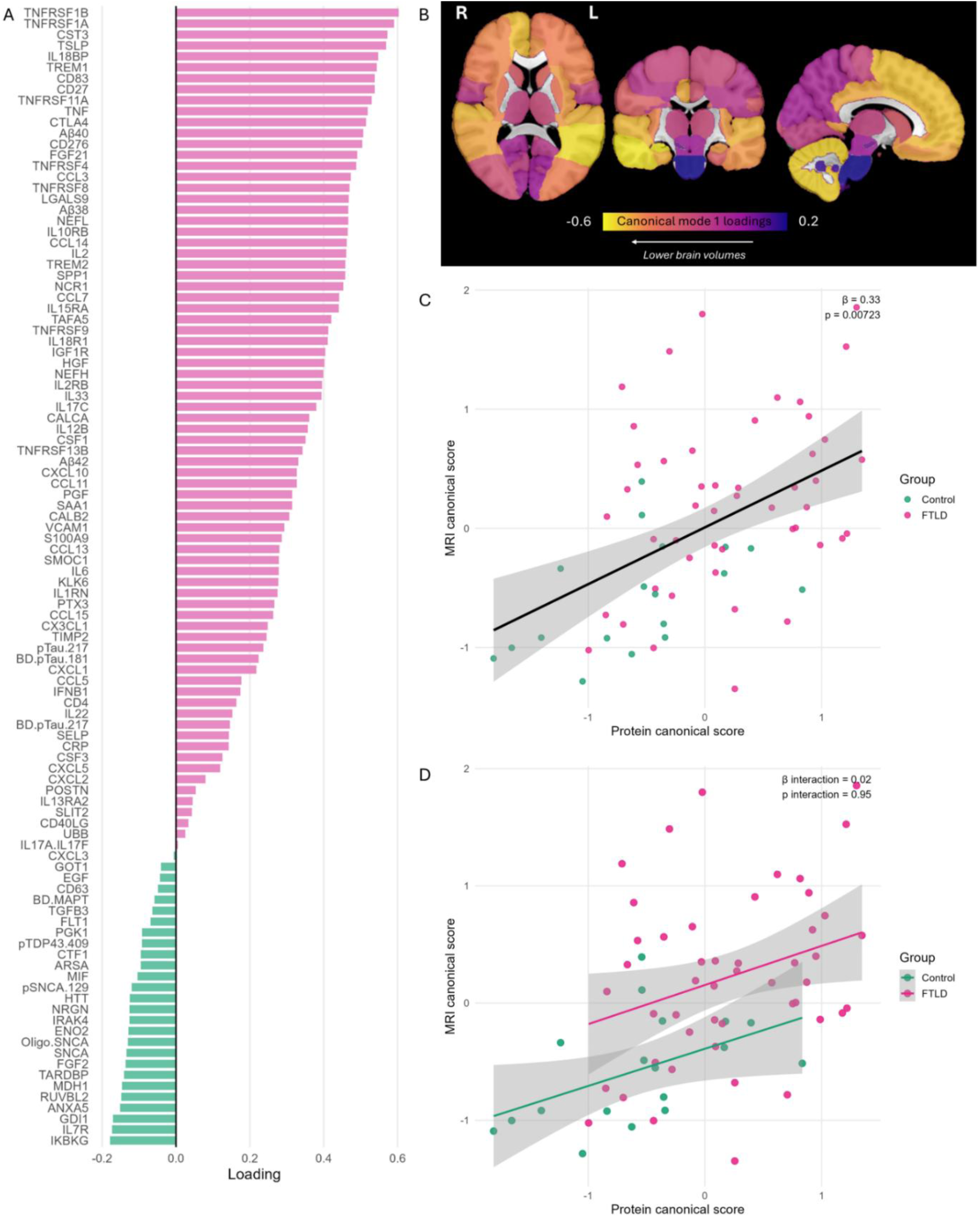
Multivariate associations between plasma protein expression and regional brain volumes. Canonical correlation analysis in participants with plasma and MRI data available within six months (n=66) identified a significant first canonical mode. Protein (A) and regional brain-volume loadings (B), and the association between participant-level protein and MRI canonical scores adjusted for group (C), are shown. Panel D shows the corresponding model including a protein-score-by-group interaction. Canonical variate signs were oriented for display; higher protein scores corresponded to higher brain-volume scores.

From the 81 brain regions, 5 had positive canonical loadings and 76 had negative loadings. The loadings of brain regions were mapped onto an image of a brain, using a colour gradient to visualise areas with higher and lower loadings (Figure 4B). The exact canonical loadings per brain region are shown in Supplementary figure 6.

A linear model was built to visualise the relationship of subject-level protein and MRI canonical scores with group (FTLD/Control) as a covariate (MRI_score ∼ Protein_score + Group) (Figure 4C). The model had a β coefficient of 0.33 and a p value of 0.0072. When an interaction term was added to the model to observe whether the relationship between MRI scores and protein scores was influenced by group, no significant interaction effect was observed (β interaction = 0.02, p value interaction = 0.95, Figure 4D).

Cox regression analysis was performed to assess associations between the protein and MRI scores of canonical mode 1 and survival in patient participants. In a model including both protein and MRI components and age and sex as covariates, the protein scores were significantly associated with survival (HR=2.56, p=0.035), whereas MRI scores were not significant (HR=0.81, p=0.62).

## Discussion

Our study demonstrates that plasma-based multiplex proteomic profiling captures extensive CNS- and inflammation-related molecular changes that are clinically relevant across the FTLD spectrum. First, CNS-related proteins distinguished participants with FTLD syndromes from healthy controls with high accuracy, driven principally by NfL, whereas inflammatory proteins provided additional information for some syndrome-versus-syndrome comparisons. Second, CD276 and SPP1 converged across differential-expression, NfL-correlation and survival analyses, linking peripheral immune signals to neuroaxonal injury and clinical outcome. Third, multivariate protein patterns enriched for immune, TNF and endothelial signalling covaried with regional brain atrophy in frontotemporal regions. Together, these findings support multiplex plasma proteomics as a means of resolving overlapping dimensions of neuronal injury and immune dysregulation in FTLD.

NfL was the most strongly increased CNS-panel protein and the best individual discriminator of FTLD syndromes from controls, consistent with its established sensitivity to neuroaxonal injury and association with progression across FTLD and other neurological disorders (25–30). In this context, NfL levels in the blood are increasingly used as a stratification tool for entry into trials, and an outcome measure for clinical trials in neurodegenerative disorders (31–33). The ten-protein CNS model improved the AUC only modestly over NfL alone (0.93 versus 0.91), suggesting that the main value of the wider panel may not be simple case-control separation. By contrast, the inflammation panel showed weaker FTLD-versus-control discrimination but stronger performance in selected syndrome comparisons, most notably PPA versus PSP. This pattern is consistent with complementary roles: a general injury marker may separate neurodegenerative disease from healthy ageing, whereas combinations of immune and tissue-response proteins may capture biological differences among clinical phenotypes. However, these models classify groups within a selected research cohort rather than establish clinical diagnostic tests. The high pairwise AUCs should be interpreted primarily from the out-of-bag bootstrap and nested cross-validation analyses, and independent validation in clinically representative populations, including diagnostic mimics, is required.

Neurogranin (NRGN) was the most significantly downregulated biomarker of the CNS panel. NRGN is a neuron specific post synaptic protein, related to synaptic plasticity, long term potentiation, and cognition (34,35). AD studies have shown that NRGN levels are significantly decreased in the blood plasma exosomes of individuals with AD and MCI but significantly increased in CSF and associated with poor cognitive outcomes (36). Post mortem studies show that NRGN is decreased in brain tissue of individuals with AD (37). There is less conclusive evidence on the role of NRGN in FTLD. Although CSF levels of NRGN were found to be increased in individuals with FTLD-induced dementia compared to FTLD-induced mild cognitive decline (MCI), no significant link with cognition was found (38).

The inflammatory profile was heterogeneous rather than uniformly pro-inflammatory. Increased proteins included TSLP, IL-6, SPP1 and several TNF-receptor-family members, alongside proteins with immunoregulatory functions such as CD276, IL-1 receptor antagonist and IL-18-binding protein (39–41). This mixture highlights the complexity and multifaceted nature of immune dysregulation in FTLD, with circulating immune proteins indexing concurrent immune activation, feedback regulation, vascular responses and tissue repair. The broad correlation of disease-associated inflammatory proteins with NfL also supports a relationship with neuroaxonal injury. CD276 (B7-H3) and SPP1 (osteopontin) were the most consistent individual candidates, being increased in FTLD and associated with both NfL and mortality. Notably, syndrome-specific analyses showed particularly pronounced upregulation in PSP and CBS, syndromes strongly associated with 4R tau pathology. CD276 is best characterised as an immune-checkpoint molecule in cancer biology, contributing to immune evasion and suppressing cytotoxic T cells and NK cells (39). Its role in neurodegeneration is largely unexplored beyond a prior report linking it to poor survival across FTD, PSP, and dementia with Lewy bodies (12). SPP1 has stronger experimental support: studies in MAPT-mutant FTD neurons and in Parkinson and Alzheimer models implicate osteopontin-related signalling in microglial recruitment, oxidative stress, synaptic engulfment and neuronal loss (42–44), supporting it as a biologically plausible and potentially targetable driver rather than a passive marker. In addition, the aforementioned NULISA study showed that SPP1 is related to survival across FTD, PSP, AD and dementia with Lewy bodies (12). The present convergence therefore nominates SPP1, and more tentatively CD276, as candidate pathways for mechanistic study and biomarker validation.

Co-expression analysis extended the individual-marker findings by identifying a module comprising immune-regulatory, endothelial and tau-related proteins (cluster 1). Cluster 1 scores were positively associated with plasma NfL, including after exclusion of NfL from the module, and did not differ significantly across FTLD-related clinical syndromes. This pattern is consistent with a shared biological signature related to neuroaxonal injury rather than a syndrome-specific profile. The module included markers of endothelial activation and vascular remodelling, including VCAM1, SLIT2, POSTN, PGF and FLT1 (45–49), alongside immune-related proteins such as CD276, CX3CL1, CSF1 and CCL11 (39,50–52), and several tau-related proteins, including pTau-217, BD-pTau-181, BD-pTau-217, and BD-MAPT. Together, these findings suggest coordinated alterations in neurovascular, immune and neurodegeneration-related pathways in FTLD (53), rather than activation of a single inflammatory mechanism. However, plasma phosphorylated-tau measures cannot be assigned to FTLD-tau pathology without molecular confirmation, particularly in the presence of possible concomitant Alzheimer pathology.

A second module (cluster 4), enriched for cellular-stress, metabolic and proteopathy-related proteins, was downregulated in FTLD and inversely related to NfL. Several of these proteins also had negative loadings in the canonical correlation analysis, implying that high levels of these markers may instead be associated with relative preservation of brain volume. The leading biomarkers of this cluster included MIF, IRAK4 and CTF1, related to cellular stress, metabolism and homeostasis, PGK1 and ANXA5, involved in metabolism and homeostasis, and TARDBP, HTT, pSNCA.129, NRGN and Oligo.SNCA, markers related to neuropathology. This internally consistent pattern may reflect relative preservation, altered release or clearance, redistribution between CNS and blood, or a compensatory response. The data do not distinguish among these possibilities, and lower plasma concentrations should not be equated with lower brain pathology. Paired plasma, cerebrospinal fluid and post-mortem tissue measurements will be needed to determine the cellular sources and direction of these changes.

Associations between individual proteins and clinical severity varied according to the clinical measure and syndrome, further supporting the context-dependent nature of peripheral immune signalling in FTLD. Higher IL-2 was associated with poorer cognitive performance on the ACE-R. As IL-2 regulates both cytotoxic and regulatory T-cell populations, this association could reflect harmful immune activation, altered immune homeostasis or a compensatory regulatory response. The latter possibility is of particular interest because low-dose IL-2 preferentially expands regulatory T cells and is being evaluated in neurodegenerative disease settings (54,55). Higher IL-18BP was associated with greater behavioural and functional impairment on the CBI-R and was also weighted positively in the protein signature associated with lower regional brain volumes. As an endogenous inhibitor of IL-18, IL-18BP may represent feedback to increased IL-18 signalling rather than a direct pathogenic process (41).This interpretation is consistent with experimental evidence that IL-18BP can attenuate neuroinflammation and cognitive impairment, whereas higher IL-18 has been associated with poorer cognition in humans (56). In participants with PSP or CBS, TNFRSF8 and PGF were associated with worse clinical severity (higher PSPRS scores), whereas FGF2, interferon-beta, TIMP2 and IL-18R1 were associated with lower scores. The TNFRSF8 finding is directionally consistent with its association with NfL and the atrophy-related protein signature, while PGF may reflect endothelial or neurovascular responses. Conversely, the associations of FGF2, interferon-beta and TIMP2 with lower severity are compatible with their reported roles in neuroinflammation, cellular stress and synaptic plasticity (57–60), but should not be interpreted as evidence of protection. The apparently contrasting associations of IL-18BP and IL-18R1 further illustrate the pleiotropic and potentially compensatory nature of IL-18 signalling, which has both inflammatory and physiological roles in the brain (61–65). Longitudinal studies should determine whether baseline protein levels predict within-person clinical decline and whether changes in these proteins track disease progression.

Interestingly, Aβ42 (amyloid-beta 42) was the only marker associated with reduced mortality risk. One possible explanation for this observation is that higher plasma Aβ42 indicates lower amyloid burden in the brain. Aβ42 is generated through cleavage of the amyloid precursor protein and constitutes one of the main components of extracellular amyloid plaques, together with Aβ40. In both CSF and plasma, lower Aβ42 levels or reduced ratios of Aβ42/Aβ40 are associated with higher amyloid deposition in the brain and are increasingly recognised as reliable fluid biomarkers of AD pathology (66). Concomitant amyloid pathology is relatively common in FTLD and has been associated with faster disease progression (67). Therefore, higher plasma Aβ42 may indicate the absence of concomitant amyloid pathology in FTLD, which would explain the reduced mortality risk.

Finally, the CCA reinforced the immune-neurodegeneration link at the brain-structural level. A protein pattern weighted towards TNF signalling, immune activation and immune regulation was associated with lower volumes across predominantly frontal, temporal and limbic regions that are vulnerable in FTLD (68–71). These findings suggest that peripheral inflammation is linked to neurodegeneration in FTLD affected brain networks, lending anatomical plausibility to the association. Together with the negative CCA loadings observed for several cluster 4 proteostasis markers, which independently correlated inversely with NfL, these results suggest a coherent pattern: pro-inflammatory signatures are associated with lower regional brain volumes, while a distinct protein set may mark relative structural preservation. This positions peripheral inflammatory proteomics as tightly coupled to and a candidate mediator of the atrophy patterns characteristic of FTLD, strengthening the case for further mechanistic study of CD276- and SPP1-related pathways. Notably, the protein component of canonical mode 1 was significantly associated with increased mortality risk, whereas the MRI component was not, suggesting that the proteomic signature may capture variation in disease progression that is not reflected by brain structural measures alone.

This study has several limitations. First, diagnoses were based on clinical criteria rather than neuropathological confirmation, raising the possibility of diagnostic misclassification. Furthermore, amyloid PET or CSF markers were not available, preventing assessment of possible Alzheimer’s disease co-pathology. This is particularly relevant given the substantial prevalence of amyloid positivity reported in FTLD syndromes, especially in older patients (72,73). Second, survival analyses were limited by the relatively small number of deaths observed (n = 40 out of 244), with most participants still alive at the time of analysis. In addition, data on disease onset were not available for many participants, therefore time to event was based on the time from the blood sample collection to the date of death or census date. Similarly limited availability of clinical assessment scores within six months of blood collections reduced the statistical power of analyses relating biomarkers to clinical function. Unequal group sizes across FTLD syndromes may also have introduced biases in pooled analyses. Stratified cross-validation and bootstrapping were used when applicable to mitigate these limitations (ROC analysis supplementary material). Third, only a portion of the cohort had MRI data available within 6 months of blood collection (n=66), limiting the statistical power of CCA. Finally, the cross-sectional observational design precludes causal inference: elevated proteins may reflect pathogenic drivers, downstream consequences, or compensatory responses, a distinction only experimental or longitudinal studies can resolve.

In conclusion, high-sensitivity plasma multiplex proteomics identified complementary CNS-related and inflammatory signatures across FTLD-related clinical syndromes. CNS-related proteins, particularly NfL, provided the strongest discrimination between participants with FTLD-related syndromes and healthy controls, whereas inflammatory proteins contributed complementary information in selected syndrome comparisons and were associated with neuroaxonal injury, regional brain volume and survival. CD276 and SPP1 emerged as prioritised candidate biomarkers through convergent associations with disease status, NfL and mortality. More broadly, the heterogeneous pattern of inflammatory, immune-regulatory and neurovascular proteins indicates that immune dysregulation in FTLD is unlikely to be represented by a single detrimental inflammatory axis. These findings support plasma proteomics as a scalable approach for biological stratification and biomarker prioritisation, rather than immediate clinical implementation. Independent, longitudinal and molecularly characterised cohorts, together with paired fluid, imaging, tissue and experimental studies, will be required to establish the temporal, cellular and mechanistic basis of the identified signatures.

## Data availability

Anonymized processed data can be shared upon request to the corresponding or senior authors. Raw data may also be requested but are likely to be subject to a data transfer agreement with restrictions required to comply with participant consent and data protection regulations.

## Acknowledgements

We thank the participant volunteers and their families for their participation in this study. We thank the National Institute for Health Research (NIHR) Cambridge Biomedical Research Centre, Cambridge University Hospitals BioRepository, and the research nurses for their contribution. We thank the and the East Anglia Dementias and Neurodegenerative Diseases Research Network (DeNDRoN) for help with subject recruitment. For the purpose of open access, the authors have applied a Creative Commons Attribution (CC BY) license to any Author Accepted Manuscript version arising from this submission.

