## Supplementary Materials for "Plasma immune signatures associate with neurodegeneration in frontotemporal lobar degeneration"

Cambridge CB2 0SZ

United Kingdom

### Supplementary methods

As an additional validation strategy for the ROC analysis, a stratified 5-fold nested cross validation was performed. Within each training fold, the same limma selection and elastic net regularisation approach was used to create the model, and performance was assessed using the test fold. ROC curves and AUC values were computed for each fold, and the pooled AUC was computed to summarise performance. Results of the ROC analysis with bootstrapping and nested cross validation are provided in supplementary figures 1 and 2 and supplementary tables 1-6).

### Supplementary Figures

**Supplementary Figure 1. Bootstrap validation of plasma protein classifiers**. Receiver-operating-characteristic curves (ROC) from 200 bootstrap samples for CNS-related and inflammation-related models distinguishing FTLD-related clinical syndromes from controls (A, B) and pairwise clinical groups (C, D). Area under the curve values and confidence intervals are shown.

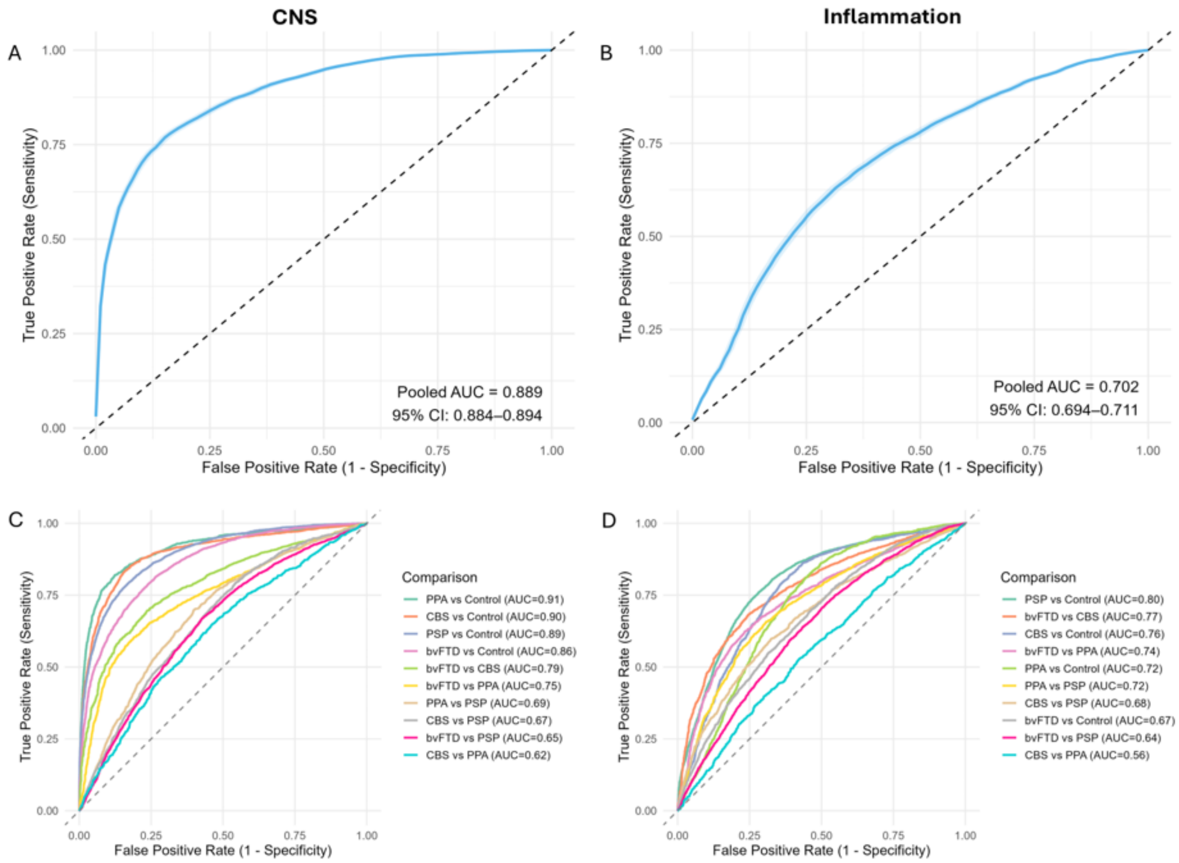

**Supplementary Figure 2. Nested cross-validation of plasma protein classifiers.**
Five-fold nested cross-validation of CNS-related and inflammation-related models distinguishing FTLD-related clinical syndromes from controls (A, B) and pairwise clinical groups (C, D). Fold-specific and pooled receiver-operating-characteristic curves are shown.

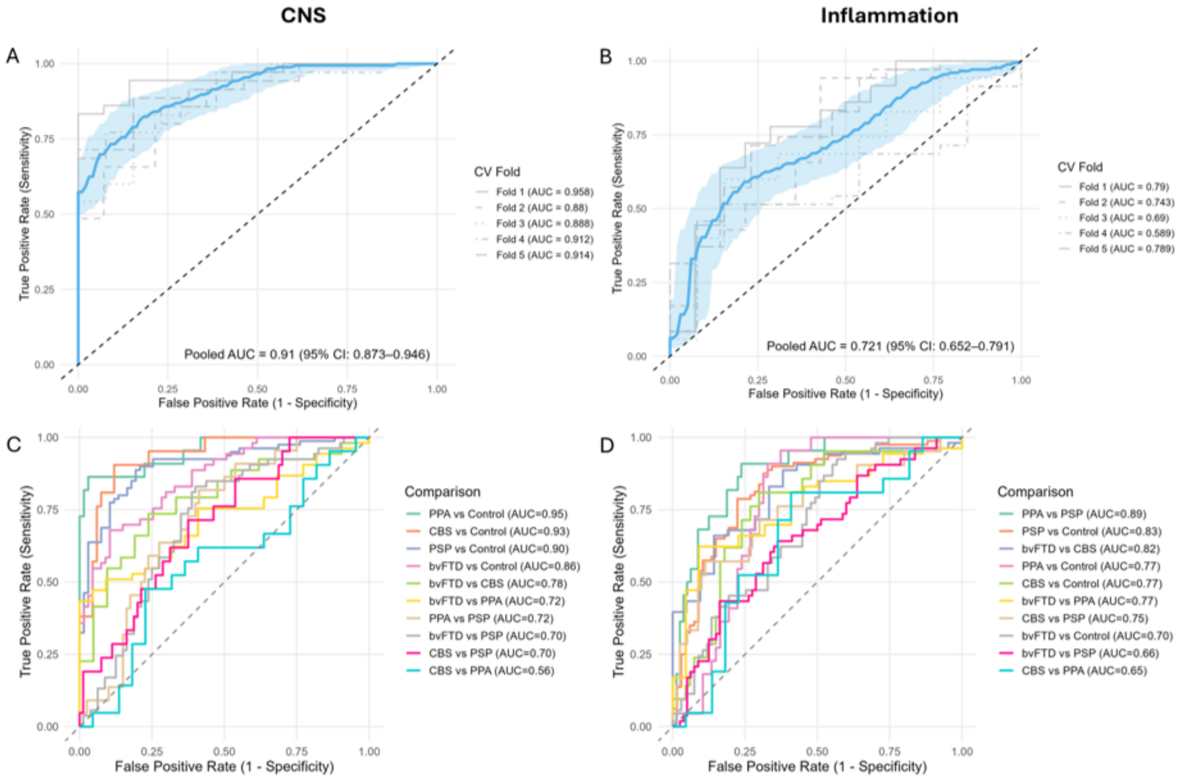

**Supplementary Figure 3. Differential expression of CNS-related plasma proteins in pairwise group comparisons.** Volcano plots from age- and sex-adjusted empirical-Bayes moderated linear models. Significant proteins were identified after false-discovery-rate correction.

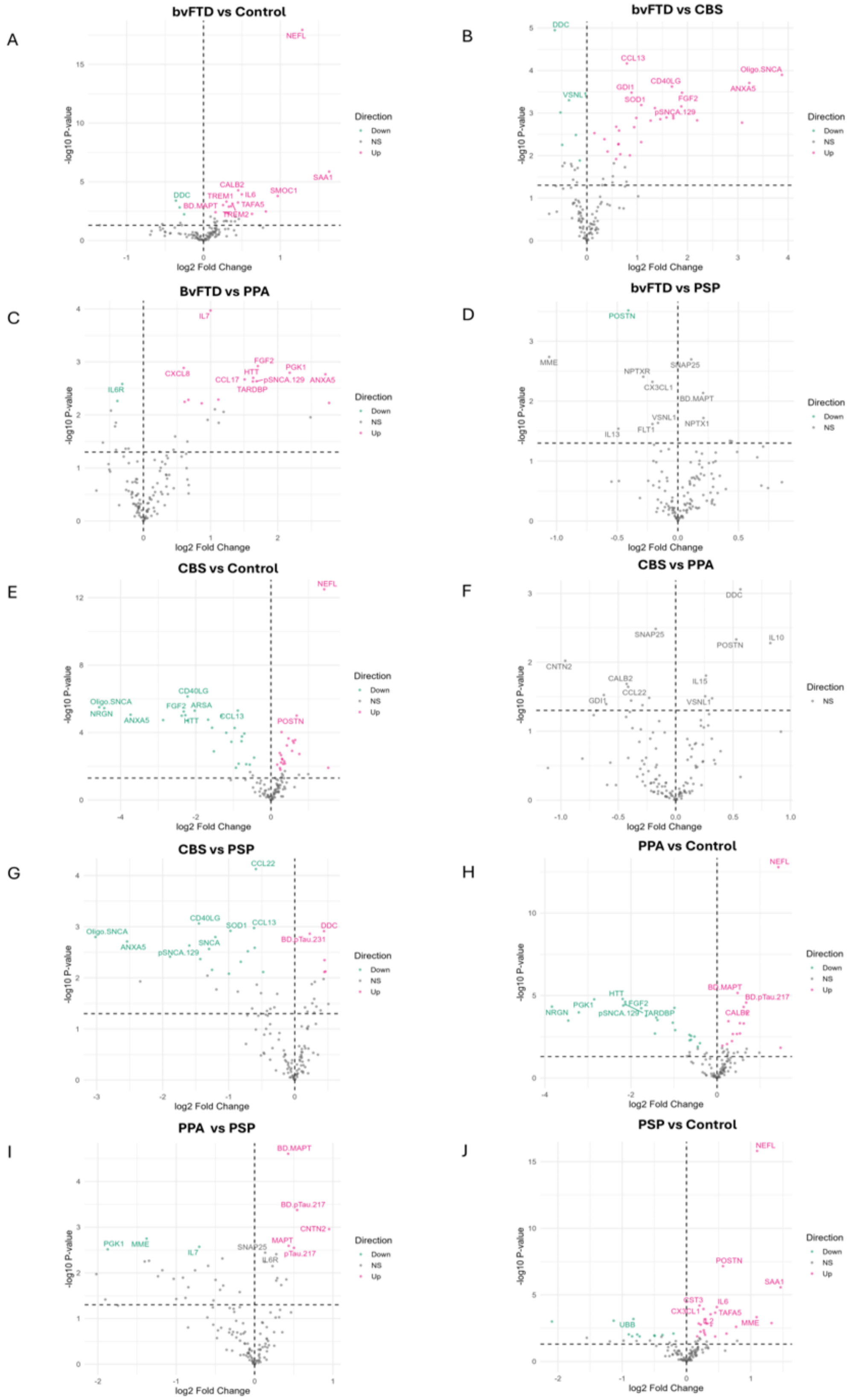

**Supplementary Figure 4. Differential expression of inflammation-related plasma proteins in pairwise group comparisons.** Volcano plots from age- and sex-adjusted empirical-Bayes moderated linear models. Significant proteins were identified after false-discovery-rate correction.

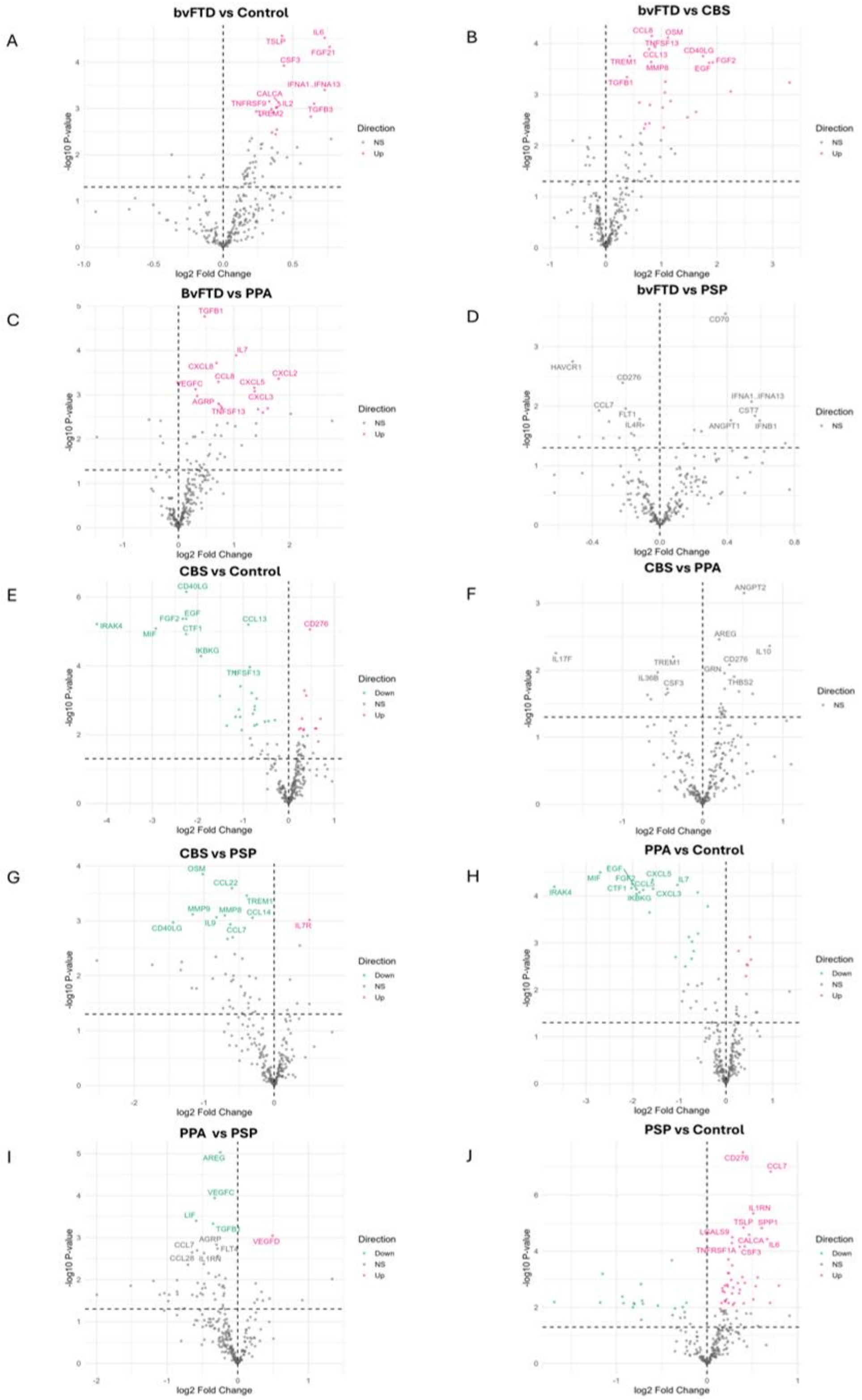

**Supplementary Figure 5. Selection and clustering of disease-associated plasma proteins.** Differentially expressed CNS-related and inflammation-related proteins are shown in A. The elbow plot used to select four clusters is shown in B, and the resulting hierarchical clustering dendrogram in C. PC1 loadings are provided in Supplementary Table 7.

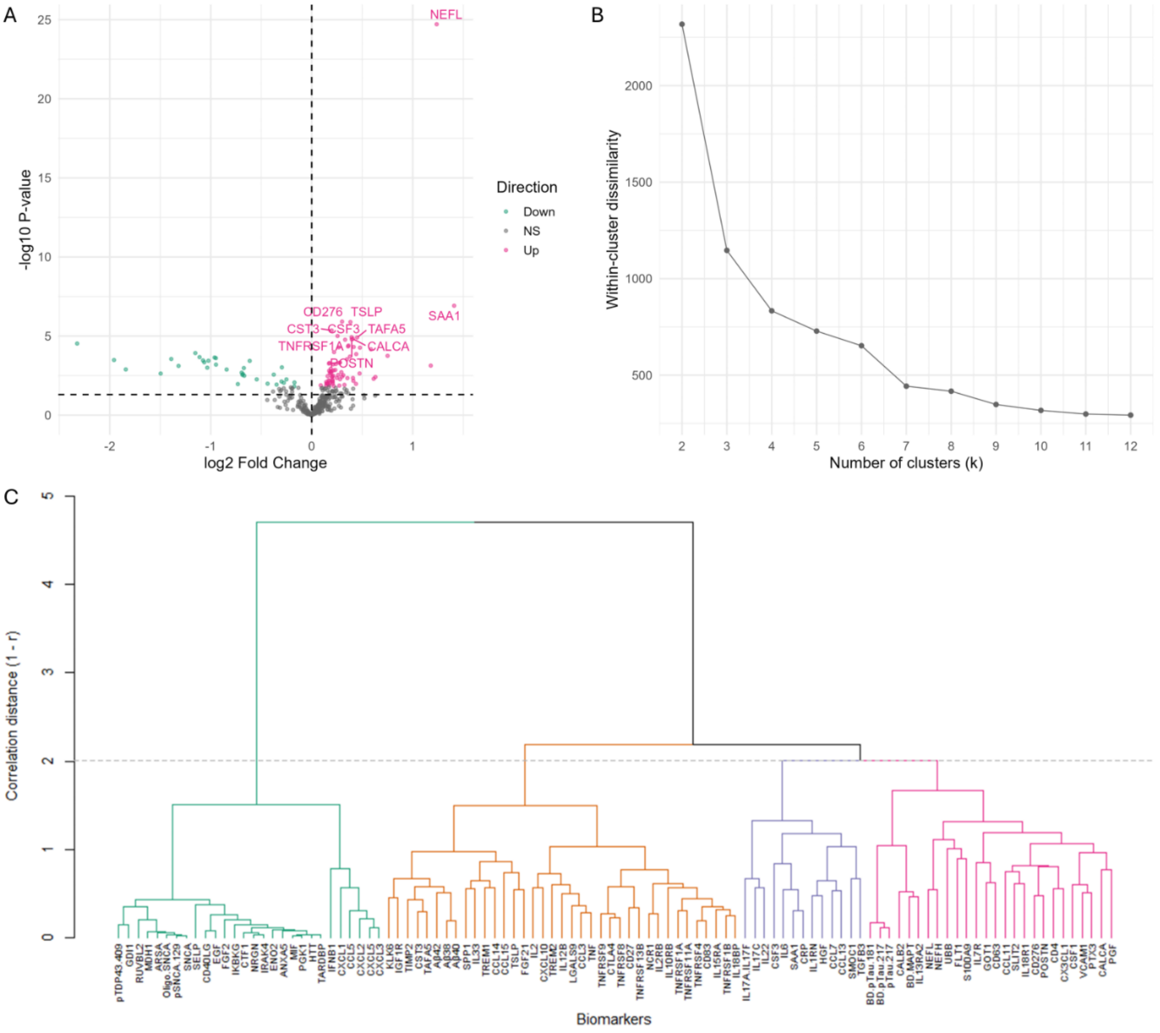

**Supplementary Figure 6. Regional brain-volume loadings from canonical correlation analysis.** Loadings of 81 regional brain volumes on the first canonical correlation mode. Direction is oriented as in Fig. 4.

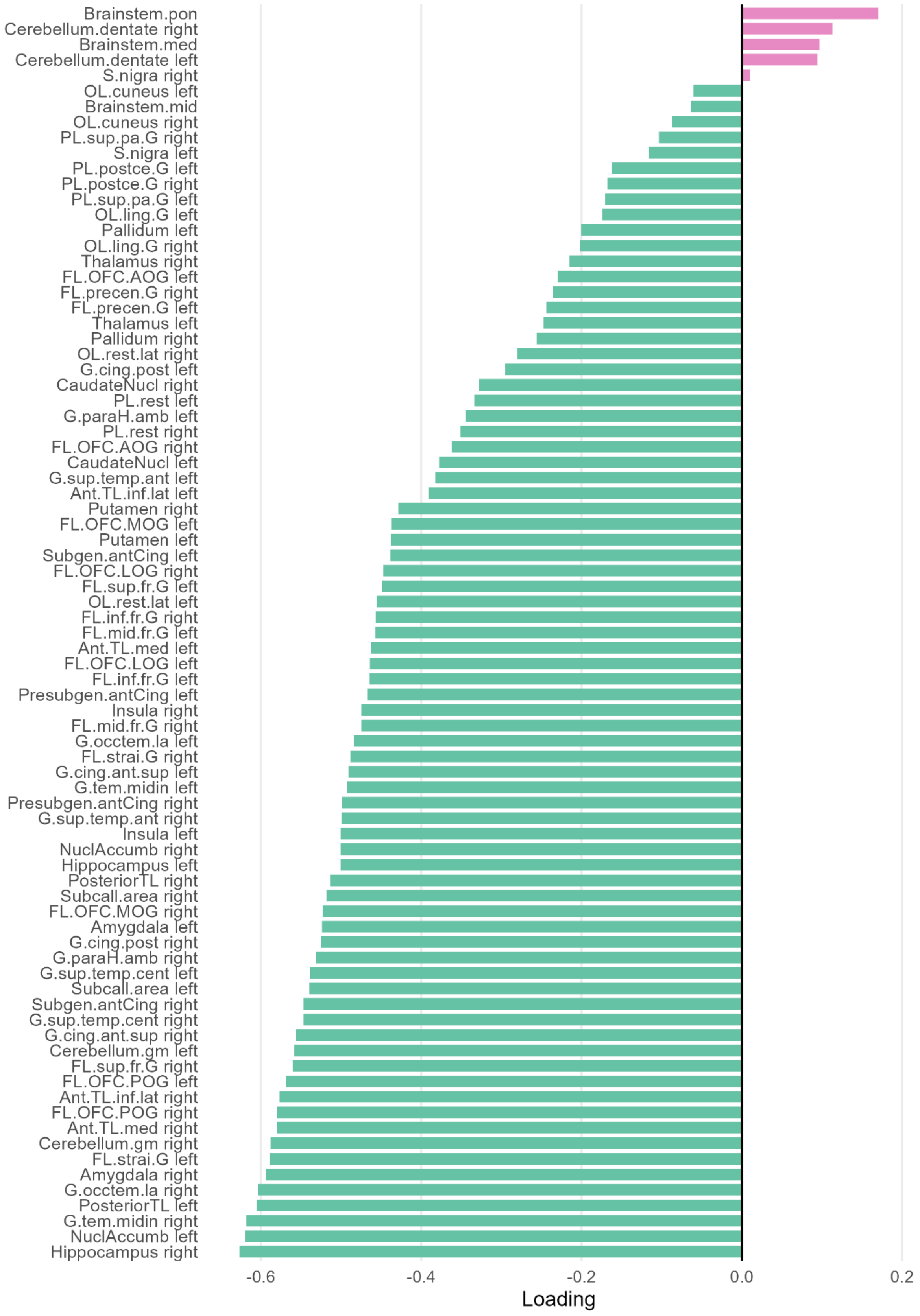

### Supplementary Tables

**Supplementary table 1:** Top 10 most frequently elastic net-selected CNS and inflammation protein biomarkers in bootstrapping ROC analysis (B = 200 bootstraps), assessing discriminatory performance between FTLD and control.

| CNS markers | | | Inflammation markers | | |
|---|---|---|---|---|---|
| Protein | Count | Frequency | Protein | Count | Frequency |
| NEFL | 200 | 1 | CD276 | 156 | 0.78 |
| SAA1 | 162 | 0.81 | TSLP | 155 | 0.775 |
| POSTN | 122 | 0.61 | IL6 | 101 | 0.505 |
| CST3 | 107 | 0.535 | TNFRSF9 | 86 | 0.43 |
| TAFA5 | 90 | 0.45 | SPP1 | 85 | 0.425 |
| NRGN | 83 | 0.415 | FGF21 | 83 | 0.415 |
| CALB2 | 71 | 0.355 | FGF2 | 61 | 0.305 |
| IL6 | 71 | 0.355 | CCL7 | 52 | 0.26 |
| IL2 | 65 | 0.325 | TNFRSF1A | 51 | 0.255 |
| SMOC1 | 64 | 0.32 | CSF3 | 46 | 0.23 |

**Supplementary table 2:** Top 10 most frequently elastic net-selected CNS markers in bootstrapping ROC analysis (B = 200 bootstraps) assessing discriminatory performance between groups.

| bvFTD vs Control | | | bvFTD vs CBS | | | bvFTD vs PPA | | | bvFTD vs PSP | | | CBS vs Control | | |
|---|---|---|---|---|---|---|---|---|---|---|---|---|---|---|
| Protein | Count | Frequency | Protein | Count | Frequency | Protein | Count | Frequency | Protein | Count | Frequency | Protein | Count | Frequency |
| NEFL | 200 | 1 | Oligo.SNCA | 170 | 0.85 | IL7 | 139 | 0.695 | POSTN | 186 | 0.93 | NEFL | 200 | 1 |
| SAA1 | 178 | 0.89 | ANXA5 | 142 | 0.71 | ANXA5 | 134 | 0.67 | NPTXR | 119 | 0.595 | CD40LG | 139 | 0.695 |
| DDC | 146 | 0.73 | DDC | 112 | 0.56 | IL6R | 109 | 0.545 | CX3CL1 | 114 | 0.57 | Oligo.SNCA | 129 | 0.645 |
| NPTXR | 93 | 0.465 | FGF2 | 112 | 0.56 | FGF2 | 105 | 0.525 | SP25 | 89 | 0.445 | NRGN | 110 | 0.55 |
| BD-MAPT | 90 | 0.45 | GDI1 | 84 | 0.42 | pSNCA.129 | 86 | 0.43 | FABP3 | 82 | 0.41 | POSTN | 85 | 0.425 |
| IL6 | 88 | 0.44 | RUVBL2 | 78 | 0.39 | PGK1 | 78 | 0.39 | MME | 76 | 0.38 | GDI1 | 79 | 0.395 |
| CALB2 | 86 | 0.43 | CCL13 | 71 | 0.355 | TARDBP | 68 | 0.34 | IL13 | 64 | 0.32 | ENO2 | 76 | 0.38 |
| IL2 | 83 | 0.415 | SOD1 | 63 | 0.315 | SFTPD | 59 | 0.295 | PGF | 50 | 0.25 | ARSA | 72 | 0.36 |
| SMOC1 | 71 | 0.355 | CXCL1 | 57 | 0.285 | pTau-217 | 50 | 0.25 | CXCL1 | 48 | 0.24 | ANXA5 | 67 | 0.335 |
| SFTPD | 61 | 0.305 | CCL26 | 55 | 0.275 | SOD1 | 48 | 0.24 | FLT1 | 44 | 0.22 | FGF2 | 63 | 0.315 |

| CBS vs PPA | | | CBS vs PSP | | | PPA vs Control | | | PPA vs PSP | | | PSP vs Control | | |
|---|---|---|---|---|---|---|---|---|---|---|---|---|---|---|
| Protein | Count | Frequency | Protein | Count | Frequency | Protein | Count | Frequency | Protein | Count | Frequency | Protein | Count | Frequency |
| GDI1 | 175 | 0.875 | CCL13 | 126 | 0.63 | NEFL | 200 | 1 | BD-MAPT | 145 | 0.725 | NEFL | 200 | 1 |
| DDC | 126 | 0.63 | CCL22 | 95 | 0.475 | HTT | 122 | 0.61 | CNTN2 | 113 | 0.565 | POSTN | 196 | 0.98 |
| IL2 | 66 | 0.33 | DDC | 95 | 0.475 | BD-MAPT | 109 | 0.545 | BD-pTau-217 | 86 | 0.43 | SAA1 | 135 | 0.675 |
| TREM1 | 66 | 0.33 | SOD1 | 92 | 0.46 | CALB2 | 85 | 0.425 | MME | 76 | 0.38 | IL6 | 91 | 0.455 |
| CNTN2 | 64 | 0.32 | CD40LG | 91 | 0.455 | IL7 | 76 | 0.38 | VEGFD | 74 | 0.37 | CST3 | 81 | 0.405 |
| SNCB | 63 | 0.315 | TREM1 | 88 | 0.44 | pTau-217 | 75 | 0.375 | PGK1 | 70 | 0.35 | PGF | 81 | 0.405 |
| SP25 | 52 | 0.26 | PSEN1 | 76 | 0.38 | FGF2 | 72 | 0.36 | HTT | 66 | 0.33 | VCAM1 | 74 | 0.37 |
| CALB2 | 51 | 0.255 | IL9 | 66 | 0.33 | BD-pTau-217 | 67 | 0.335 | IL7 | 66 | 0.33 | TAFA5 | 67 | 0.335 |
| IL15 | 51 | 0.255 | Oligo.SNCA | 64 | 0.32 | NRGN | 65 | 0.325 | UCHL1 | 64 | 0.32 | CXCL10 | 63 | 0.315 |
| POSTN | 50 | 0.25 | SNCA | 63 | 0.315 | IL2 | 54 | | SQSTM1 | 63 | 0.315 | IL2 | 60 | 0.3 |

**Supplementary table 3:** Top 10 most frequently elastic net-selected inflammation markers in bootstrapping ROC analysis (B = 200 bootstraps) assessing discriminatory performance between groups.

| bvFTD vs Control | | | bvFTD vs CBS | | | bvFTD vs PPA | | | bvFTD vs PSP | | | CBS vs Control | | |
|---|---|---|---|---|---|---|---|---|---|---|---|---|---|---|
| Protein | Count | Frequency | Protein | Count | Frequency | Protein | Count | Frequency | Protein | Count | Frequency | Protein | Count | Frequency |
| CSF3 | 181 | 0.905 | FGF2 | 123 | 0.615 | IL7 | 110 | 0.55 | HAVCR1 | 179 | 0.895 | CD40LG | 150 | 0.75 |
| IFNA1/IFNA13 | 110 | 0.55 | MMP8 | 104 | 0.52 | MIF | 105 | 0.525 | CD276 | 162 | 0.81 | IRAK4 | 126 | 0.63 |
| TSLP | 109 | 0.545 | IRAK4 | 88 | 0.44 | CXCL2 | 104 | 0.52 | IL15RA | 102 | 0.51 | FGF2 | 125 | 0.625 |
| IL1RN | 81 | 0.405 | TNFSF13 | 85 | 0.425 | TGFB1 | 95 | 0.475 | CCL7 | 85 | 0.425 | CD276 | 117 | 0.585 |
| FGF21 | 75 | 0.375 | FGF23 | 80 | 0.4 | CD93 | 91 | 0.455 | OSMR | 77 | 0.385 | EGF | 93 | 0.465 |
| IL17C | 74 | 0.37 | OSM | 74 | 0.37 | IL6R | 87 | 0.435 | IFNB1 | 72 | 0.36 | CCL13 | 81 | 0.405 |
| IL6 | 70 | 0.35 | MIF | 73 | 0.365 | FGF2 | 85 | 0.425 | FLT1 | 51 | 0.255 | MIF | 81 | 0.405 |
| CALCA | 69 | 0.345 | CXCL1 | 64 | 0.32 | IRAK4 | 69 | 0.345 | CST7 | 48 | 0.24 | CTF1 | 68 | 0.34 |
| IL2 | 63 | 0.315 | CCL26 | 57 | 0.285 | CXCL3 | 63 | 0.315 | AREG | 45 | 0.225 | MMP8 | 50 | 0.25 |
| TNFRSF9 | 45 | 0.225 | CD40LG | 56 | 0.28 | CXCL5 | 54 | 0.27 | CD70 | 45 | 0.225 | IKBKG | 48 | 0.24 |

| CBS vs PPA | | | CBS vs PSP | | | PPA vs Control | | | PPA vs PSP | | | PSP vs Control | | |
|---|---|---|---|---|---|---|---|---|---|---|---|---|---|---|
| Protein | Count | Frequency | Protein | Count | Frequency | Protein | Count | Frequency | Protein | Count | Frequency | Protein | Count | Frequency |
| IL36G | 99 | 0.495 | TREM1 | 140 | 0.7 | CCL5 | 110 | 0.55 | AREG | 148 | 0.74 | CD276 | 199 | 0.995 |
| IL36B | 91 | 0.455 | CCL14 | 109 | 0.545 | MIF | 95 | 0.475 | VEGFD | 131 | 0.655 | CCL7 | 167 | 0.835 |
| TREM1 | 80 | 0.4 | MMP8 | 109 | 0.545 | CXCL3 | 79 | 0.395 | TGFB1 | 125 | 0.625 | SPP1 | 159 | 0.795 |
| AREG | 75 | 0.375 | IL7R | 101 | 0.505 | CXCL2 | 77 | 0.385 | LIF | 92 | 0.46 | TSLP | 118 | 0.59 |
| ANGPT2 | 73 | 0.365 | CCL13 | 83 | 0.415 | IRAK4 | 76 | 0.38 | AGRP | 87 | 0.435 | IL6 | 93 | 0.465 |
| GRN | 60 | 0.3 | MMP9 | 81 | 0.405 | FGF2 | 75 | 0.375 | FLT4 | 86 | 0.43 | IL1RN | 77 | 0.385 |
| THBS2 | 53 | 0.265 | CD40LG | 76 | 0.38 | CXCL5 | 74 | 0.37 | VEGFC | 64 | 0.32 | CALCA | 66 | 0.33 |
| CSF2 | 47 | 0.235 | CCL22 | 74 | 0.37 | SELP | 66 | 0.33 | PGF | 54 | 0.27 | CCL14 | 57 | 0.285 |
| VEGFD | 47 | 0.235 | OSM | 74 | 0.37 | CTF1 | 65 | 0.325 | IL5RA | 47 | 0.235 | TNFRSF1A | 56 | 0.28 |
| FLT4 | 45 | 0.225 | IL9 | 64 | 0.32 | IKBKG | 64 | 0.32 | IL13RA2 | 44 | 0.22 | LGALS9 | 45 | 0.225 |

**Supplementary table 4:** Top 10 most frequently elastic net-selected CNS and inflammation markers in 5-fold nested cross-validation ROC analysis, assessing discriminatory performance between FTLD and control.

| CNS markers | | | Inflammation markers | | |
|---|---|---|---|---|---|
| Protein | Count | Frequency | Protein | Count | Frequency |
| NEFL | 5 | 1 | CD276 | 5 | 1 |
| SAA1 | 5 | 1 | TSLP | 5 | 1 |
| IL2 | 4 | 0.8 | IL6 | 4 | 0.8 |
| POSTN | 4 | 0.8 | SPP1 | 4 | 0.8 |
| IL6 | 3 | 0.6 | TNFRSF9 | 4 | 0.8 |
| NRGN | 3 | 0.6 | CTF1 | 3 | 0.6 |
| TAFA5 | 3 | 0.6 | CALCA | 2 | 0.4 |
| CALB2 | 2 | 0.4 | CCL7 | 2 | 0.4 |
| CST3 | 2 | 0.4 | CSF3 | 2 | 0.4 |
| SMOC1 | 2 | 0.4 | LGALS9 | 2 | 0.4 |

**Supplementary table 5:** Top 10 most frequently elastic net-selected CNS proteins in 5-fold nested cross-validation ROC analysis, assessing discriminatory performance between groups.

| bvFTD vs Control | | | bvFTD vs CBS | | | bvFTD vs PPA | | | bvFTD vs PSP | | | CBS vs Control | | |
|---|---|---|---|---|---|---|---|---|---|---|---|---|---|---|
| Protein | Count | Frequency | Protein | Count | Frequency | Protein | Count | Frequency | Protein | Count | Frequency | Protein | Count | Frequency |
| DDC | 5 | 1 | Oligo.SNCA | 5 | 1 | pSNCA.129 | 5 | 1 | NPTXR | 5 | 1 | NEFL | 5 | 1 |
| NEFL | 5 | 1 | ANXA5 | 4 | 0.8 | ANXA5 | 4 | 0.8 | POSTN | 5 | 1 | Oligo.SNCA | 5 | 1 |
| SAA1 | 5 | 1 | DDC | 4 | 0.8 | IL7 | 4 | 0.8 | MME | 4 | 0.8 | CD40LG | 3 | 0.6 |
| BD-MAPT | 3 | 0.6 | GDI1 | 4 | 0.8 | FGF2 | 3 | 0.6 | CX3CL1 | 3 | 0.6 | NRGN | 3 | 0.6 |
| CALB2 | 3 | 0.6 | CCL13 | 3 | 0.6 | IL6R | 2 | 0.4 | FABP3 | 3 | 0.6 | ARSA | 2 | 0.4 |
| CRP | 2 | 0.4 | FGF2 | 3 | 0.6 | Oligo.SNCA | 2 | 0.4 | PGF | 3 | 0.6 | CCL13 | 2 | 0.4 |
| IL2 | 2 | 0.4 | RUVBL2 | 3 | 0.6 | PGK1 | 2 | 0.4 | SP25 | 3 | 0.6 | GDI1 | 2 | 0.4 |
| IL6 | 2 | 0.4 | ENO2 | 2 | 0.4 | pTau-217 | 2 | 0.4 | BD-MAPT | 2 | 0.4 | HTT | 2 | 0.4 |
| NPTXR | 2 | 0.4 | POSTN | 2 | 0.4 | TARDBP | 2 | 0.4 | IL13 | 2 | 0.4 | ANXA5 | 1 | 0.2 |
| SFTPD | 2 | 0.4 | SOD1 | 2 | 0.4 | CCL17 | 1 | 0.2 | CCL11 | 1 | 0.2 | ENO2 | 1 | 0.2 |
| CBS vs PPA | | | CBS vs PSP | | | PPA vs Control | | | PPA vs PSP | | | PSP vs Control | | |
| Protein | Count | Frequency | Protein | Count | Frequency | Protein | Count | Frequency | Protein | Count | Frequency | Protein | Count | Frequency |
| DDC | 5 | 1 | CCL13 | 4 | 0.8 | NEFL | 5 | 1 | BD-MAPT | 5 | 1 | NEFL | 5 | 1 |
| GDI1 | 5 | 1 | CD40LG | 4 | 0.8 | BD-MAPT | 4 | 0.8 | BD-pTau-217 | 4 | 0.8 | POSTN | 5 | 1 |
| IL2 | 4 | 0.8 | pSNCA.129 | 4 | 0.8 | HTT | 4 | 0.8 | CNTN2 | 4 | 0.8 | SAA1 | 4 | 0.8 |
| CCL22 | 2 | 0.4 | SOD1 | 4 | 0.8 | NRGN | 4 | 0.8 | HTT | 3 | 0.6 | CST3 | 3 | 0.6 |
| CNTN2 | 2 | 0.4 | ANXA5 | 3 | 0.6 | pTau-217 | 3 | 0.6 | PGK1 | 3 | 0.6 | CXCL10 | 3 | 0.6 |
| ENO2 | 2 | 0.4 | ARSA | 3 | 0.6 | ANXA5 | 2 | 0.4 | SQSTM1 | 3 | 0.6 | VCAM1 | 3 | 0.6 |
| IL15 | 2 | 0.4 | CCL22 | 3 | 0.6 | CD40LG | 2 | 0.4 | IL6R | 2 | 0.4 | IL6 | 2 | 0.4 |
| MME | 2 | 0.4 | DDC | 3 | 0.6 | IL2 | 2 | 0.4 | IL7 | 2 | 0.4 | CCL11 | 1 | 0.2 |
| Oligo.SNCA | 2 | 0.4 | HTT | 3 | 0.6 | ARSA | 1 | 0.2 | MME | 2 | 0.4 | CRP | 1 | 0.2 |
| SNCB | 2 | 0.4 | Oligo.SNCA | 3 | 0.6 | CALB2 | 1 | 0.2 | pSNCA.129 | 2 | 0.4 | FGF2 | 1 | 0.2 |

**Supplementary table 6:** Top 10 most frequently elastic net-selected inflammation proteins in 5-fold nested cross-validation ROC analysis, assessing discriminatory performance between groups.

| bvFTD vs Control | | | bvFTD vs CBS | | | bvFTD vs PPA | | | bvFTD vs PSP | | | CBS vs Control | | |
|---|---|---|---|---|---|---|---|---|---|---|---|---|---|---|
| Protein | Count | Frequency | Protein | Count | Frequency | Protein | Count | Frequency | Protein | Count | Frequency | Protein | Count | Frequency |
| CSF3 | 5 | 1 | FGF2 | 4 | 0.8 | IL7 | 5 | 1 | CD276 | 5 | 1 | FGF2 | 5 | 1 |
| FGF21 | 4 | 0.8 | IRAK4 | 4 | 0.8 | MIF | 4 | 0.8 | HAVCR1 | 5 | 1 | CD276 | 4 | 0.8 |
| IFNA1/IFNA13 | 4 | 0.8 | TNFSF13 | 4 | 0.8 | CD93 | 3 | 0.6 | IL15RA | 4 | 0.8 | CD40LG | 4 | 0.8 |
| IL1RN | 4 | 0.8 | FGF23 | 3 | 0.6 | CXCL2 | 3 | 0.6 | CCL7 | 3 | 0.6 | IKBKG | 3 | 0.6 |
| IL17C | 3 | 0.6 | MMP8 | 3 | 0.6 | FGF2 | 3 | 0.6 | AREG | 2 | 0.4 | IRAK4 | 3 | 0.6 |
| IL2 | 3 | 0.6 | CCL26 | 2 | 0.4 | IL6R | 3 | 0.6 | FGF23 | 2 | 0.4 | MIF | 3 | 0.6 |
| TSLP | 3 | 0.6 | CXCL1 | 2 | 0.4 | TGFB1 | 3 | 0.6 | IFNB1 | 2 | 0.4 | CCL13 | 2 | 0.4 |
| CALCA | 2 | 0.4 | MIF | 2 | 0.4 | AGRP | 1 | 0.2 | IL7R | 2 | 0.4 | CCL5 | 2 | 0.4 |
| IL6 | 2 | 0.4 | OSM | 2 | 0.4 | ANGPT1 | 1 | 0.2 | TNFSF15 | 2 | 0.4 | EGF | 2 | 0.4 |
| CCL8 | 1 | 0.2 | CCL13 | 1 | 0.2 | CCL5 | 1 | 0.2 | ANGPT1 | 1 | 0.2 | CTF1 | 1 | 0.2 |

| CBS vs PPA | | | CBS vs PSP | | | PPA vs Control | | | PPA vs PSP | | | PSP vs Control | | |
|---|---|---|---|---|---|---|---|---|---|---|---|---|---|---|
| Protein | Count | Frequency | Protein | Count | Frequency | Protein | Count | Frequency | Protein | Count | Frequency | Protein | Count | Frequency |
| ANGPT2 | 4 | 0.8 | MMP8 | 5 | 1 | CXCL2 | 5 | 1 | AREG | 5 | 1 | CCL7 | 5 | 1 |
| GRN | 4 | 0.8 | TREM1 | 5 | 1 | CCL5 | 3 | 0.6 | TGFB1 | 5 | 1 | CD276 | 5 | 1 |
| IL36G | 4 | 0.8 | CCL14 | 4 | 0.8 | CTF1 | 3 | 0.6 | VEGFD | 5 | 1 | SPP1 | 5 | 1 |
| TREM1 | 4 | 0.8 | CD40LG | 4 | 0.8 | MIF | 3 | 0.6 | AGRP | 4 | 0.8 | IL6 | 4 | 0.8 |
| AREG | 3 | 0.6 | IL7R | 4 | 0.8 | CXCL3 | 2 | 0.4 | FLT4 | 4 | 0.8 | TSLP | 4 | 0.8 |
| IL36B | 3 | 0.6 | CCL22 | 3 | 0.6 | CXCL5 | 2 | 0.4 | LIF | 4 | 0.8 | CCL14 | 3 | 0.6 |
| CCL22 | 2 | 0.4 | IL9 | 3 | 0.6 | IKBKG | 2 | 0.4 | VEGFC | 3 | 0.6 | IL1RN | 3 | 0.6 |
| FLT4 | 2 | 0.4 | OSM | 3 | 0.6 | IL1B | 2 | 0.4 | IL13RA2 | 2 | 0.4 | CALCA | 2 | 0.4 |
| IL17F | 2 | 0.4 | CCL13 | 2 | 0.4 | IRAK4 | 2 | 0.4 | PGF | 2 | 0.4 | LGALS9 | 2 | 0.4 |
| THBS2 | 2 | 0.4 | FLT3LG | 2 | 0.4 | CSF3 | 1 | 0.2 | CCL28 | 1 | 0.2 | TNFRSF1A | 2 | 0.4 |

**Supplementary table 7:** Principal component 1 (PC1) loadings of biomarker clusters computed by performing hierarchical clustering on differentially expressed CNS and inflammation biomarkers, followed by independent PCA on each cluster.

| Cluster 1 | | Cluster 2 | | Cluster 3 | | Cluster 4 | |
|---|---|---|---|---|---|---|---|
| Biomarker | Loading | Biomarker | Loading | Biomarker | Loading | Biomarker | Loading |
| VCAM1 | 0.740225 | SAA1 | 0.717378 | TNFRSF1B | 0.860202 | MIF | 0.977973 |
| pTau.217 | 0.681979 | HGF | 0.714725 | TNFRSF4 | 0.818124 | PGK1 | 0.976288 |
| SLIT2 | 0.61995 | IL1RN | 0.7039 | CD83 | 0.816389 | TARDBP | 0.97253 |
| BD.pTau.181 | 0.615698 | IL6 | 0.665629 | TNFRSF11A | 0.813963 | HTT | 0.972029 |
| CD276 | 0.603106 | CRP | 0.63089 | TNF | 0.799929 | pSNCA.129 | 0.967889 |
| CX3CL1 | 0.592402 | CCL7 | 0.609764 | IL15RA | 0.799298 | CTF1 | 0.963196 |
| BD.pTau.217 | 0.56409 | CCL13 | 0.475219 | TNFRSF1A | 0.797085 | IRAK4 | 0.962405 |
| IL13RA2 | 0.523421 | CSF3 | 0.426117 | LGALS9 | 0.772931 | NRGN | 0.95052 |
| POSTN | 0.495095 | SMOC1 | 0.422616 | CTLA4 | 0.772106 | ANXA5 | 0.947444 |
| CCL11 | 0.47672 | IL17C | 0.4027 | CST3 | 0.768801 | Oligo.SNCA | 0.94416 |
| IL18R1 | 0.473327 | IL17A.IL17F | 0.397071 | IL18BP | 0.762293 | ENO2 | 0.9382 |
| CSF1 | 0.461358 | TGFB3 | 0.363608 | CD27 | 0.742696 | ARSA | 0.933611 |
| PGF | 0.458243 | IL22 | 0.336173 | TNFRSF9 | 0.731798 | SNCA | 0.931966 |
| BD.MAPT | 0.438163 | | | TNFRSF8 | 0.730223 | MDH1 | 0.919452 |
| CD4 | 0.422624 | | | Aβ40 | 0.720208 | EGF | 0.916721 |
| CALB2 | 0.418861 | | | CCL3 | 0.716313 | IKBKG | 0.89267 |
| NEFL | 0.38015 | | | IL2RB | 0.703548 | CD40LG | 0.881965 |
| PTX3 | 0.359155 | | | TIMP2 | 0.697537 | pTDP43.409 | 0.876957 |
| NEFH | 0.335716 | | | TAFA5 | 0.693583 | FGF2 | 0.874516 |
| IL7R | 0.32749 | | | NCR1 | 0.67198 | GDI1 | 0.818542 |
| GOT1 | 0.322751 | | | IL33 | 0.651616 | SELP | 0.78212 |
| CD63 | 0.316144 | | | IL10RB | 0.640197 | RUVBL2 | 0.778878 |
| FLT1 | 0.289408 | | | TSLP | 0.617524 | CXCL3 | 0.659781 |
| CALCA | 0.281586 | | | IL12B | 0.604493 | CXCL2 | 0.647136 |
| UBB | -0.04817 | | | TNFRSF13B | 0.600926 | CXCL5 | 0.626746 |
| S100A9 | -0.00615 | | | Aβ38 | 0.577118 | CCL5 | 0.467894 |
| | | | | TREM1 | 0.574785 | IFNB1 | 0.400171 |
| | | | | Aβ42 | 0.566768 | CXCL1 | 0.366862 |
| | | | | CCL14 | 0.551953 | | |
| | | | | TREM2 | 0.551368 | | |
| | | | | SPP1 | 0.512885 | | |
| | | | | CXCL10 | 0.507355 | | |
| | | | | FGF21 | 0.499389 | | |
| | | | | KLK6 | 0.490076 | | |
| | | | | IGF1R | 0.48314 | | |
| | | | | IL2 | 0.475427 | | |
| | | | | CCL15 | 0.431027 | | |
